# The Calculus of Confidence: Modelling Vaccine Hesitancy and Strategies for Support

**DOI:** 10.64898/2025.12.21.25342784

**Authors:** Erin E. Gill, Geoffrey L. Winsor, Baofeng Jia, Justin Cook, Larisa Lotoski, Maria V. Medeleanu, Erica Di Ruggiero, Emily Cameron, Marc-André Langois, Theo J. Moraes, Elinor Simons, Padmaja Subbarao, Stuart E. Turvey, Meghan B. Azad, Fiona S.L. Brinkman

## Abstract

**Background:** Vaccine hesitancy is a growing issue that the WHO ranks as one of the top 10 threats to global health. Public confidence in vaccines and rates of routine childhood vaccination have been declining around the world since the pandemic, when many countries saw the instatement of COVID-19 vaccine mandates.

**Objectives:** We leveraged the COVID-19 add-on study, conducted by the CHILD Cohort Study (Canada’s most phenotypically diverse, large prospective longitudinal birth cohort), to determine characteristics associated with adult participants’ vaccine hesitancy. Our goal was to identify potential strategies for addressing vaccine uptake concerns. This study complements others by examining more behavioural, socioeconomic, attitudinal and additional characteristics, in some cases with greater granularity, and by further exploring the effects of COVID-19 vaccine mandates on vaccine uptake and beliefs.

**Methods:** We generated penalized logistic regression models and used statistical tests to analyze a dataset of nearly 700 questionnaire responses where vaccine hesitancy was measured by participants’ agreement or disagreement with the following statements: “Getting myself vaccinated is important for the health of others in my community” and “Getting vaccinated is a good way to protect myself from disease’’. We also examined whether vaccination status or opinions changed after the imposition of vaccine mandates.

**Results:** While vaccine mandates were successful in increasing COVID-19 specific vaccine uptake in hesitant individuals vs confident individuals, they were ineffective in modifying hesitant individuals’ beliefs about vaccines. Vaccine-confident individuals were more likely to engage in pandemic safety measures such as physical distancing, while vaccine-hesitant individuals were more likely to have or had chronic medical conditions in the past, experience economic precarity, have lower socioeconomic status and/or formal education level, have had difficulty accessing medical care, and rely on friends and internet sites that were not governmental for COVID-19 related information.

**Conclusions:** To ensure that relevant information regarding vaccines reaches all segments of the population, outreach strategies should be tailored to individuals with a variety of cultural or educational backgrounds. Improving access to medical care could also improve access to reliable information. Vaccine mandates do not impact an individual’s beliefs in vaccines, and so countering vaccine hesitancy itself is likely to be more effective in terms of ensuring continuous vaccine uptake in a population.

## Introduction

Vaccines are one of public health’s most cost-effective interventions to control and prevent disease in populations. Many countries have led widespread public vaccination campaigns and have experienced concomitant declines in disease rates to the extent that certain viral and bacterial illnesses have been nearly or completely eradicated [1]. The average life expectancy in such countries has risen by decades over the last 100 years. A recent study suggests that 40% of the drastic decline in global infant mortality over the last 50 years is due to vaccine administration [2].

The WHO Strategic Advisory Group of Experts (SAGE) on Vaccine Hesitancy define vaccine hesitancy as a “delay in acceptance or refusal of vaccination despite availability of vaccination services”, and stress that hesitancy exists on a continuum from complete refusal to partial acceptance [3]. The SAGE concluded that in order for vaccine confidence to exist, one must have three-fold trust in 1) vaccine safety and effectiveness, 2) the healthcare system administering vaccines and 3) the individuals making the policies that determine which vaccines are necessary [3]. Many elements can influence vaccine hesitancy including one’s history, socioeconomic status, political and personal beliefs.

While vaccine hesitancy has existed since vaccination began, it is now considered one of the top 10 threats to global health by the WHO [4]. There has been a recent rise in anti-vaccine sentiment both in Canada and globally [5] - Canada has lost its measles-free designation (there have been > 5,000 cases in 2025 [6]), and the United States could soon follow [7]. Many attitudinal shifts have occurred over the last decade, a time period that coincides with the COVID-19 pandemic and vaccination mandates in some locations [8]. Public confidence in vaccine effectiveness and safety saw a large decrease over the course of the pandemic in Canada in young adults, particularly those ages 18-24 (Figure S1; [9]), while confidence increased for those over 55 (but still declined overall since 2015). These trends are not unique to Canada as they are being seen throughout Europe [10] and the WHO-UNICEF Estimates of National Immunization Coverage have shown a decrease in uptake for many routine childhood vaccines throughout South America, Central America, Asia and the USA since the pandemic [10]. Critically, vaccine mandates led to individuals in many countries becoming vaccinated for COVID-19 during the pandemic (effects were dependent on vaccine supply and pre-mandate vaccine uptake level within the population) [11,12], but subsequent backlash has since led to reduced levels of routine childhood vaccines [13] and fostered increased skepticism of vaccine effectiveness [14]. The pandemic saw both successes and failures in the implementation of such mandates [13], but learning about concepts such as vaccine effectiveness and herd immunity can lead to increased intentions to vaccinate [14,15]. Unfortunately, vulnerable populations are now facing increased morbidity and mortality due to the growing gap in routine vaccine coverage [17].

We leveraged the COVID-19 add-on study [18] of the CHILD cohort [19,20] to model adult participants’ general vaccine beliefs during the pandemic, as gauged by their agreement or disagreement with the statements **“Getting myself vaccinated is important for the health of others in my community”** (“community” statement) and **“Getting vaccinated is a good way to protect myself from disease’’** (“self” statement). We also examined how individuals’ beliefs about vaccination are related to their household income, education, where they chose to obtain COVID-19-related information and their decisions to receive vaccines (COVID-19 vaccine, influenza vaccine or both). In addition, this study aimed to determine how individuals’ opinions towards vaccination changed following the imposition of vaccine mandates. While literature exists describing correlations between individuals’ opinions regarding vaccines and other characteristics [21–25] or their vaccination status and other characteristics [26,27], there have been few investigations into whether opinions about vaccines correlate with vaccination status. We investigated whether vaccination rates or changes in vaccination status following vaccine mandates were associated with vaccine beliefs. Our analyses have identified key variables associated with vaccine hesitancy of Canadians that could be used to design improvements to public health campaigns involving vaccination. It may be more effective to design approaches more directly addressing vaccine hesitancy, rather than mandating vaccination only during epidemics or pandemics. This could better foster public trust in science, and help individuals build habits that are long-lasting - improving their health and public health overall.

## Methods

### Data Collection

Data were collected from the CHILD Cohort Study [7,8], involving 3,454 healthy Canadian infant participants and their families. CHILD is based around four hub cities across the country (Edmonton, Toronto, Vancouver and Winnipeg) and ethics were approved at the University of Alberta, McMaster University, the University of British Columbia, the University of Manitoba and the Hospital for Sick Children in Toronto. The study data were collected and managed using REDCap (Research Electronic Data Capture) hosted at the British Columbia Children’s Hospital Research Institute (BCCHR) [28,29]. REDCap is a secure, web-based software platform designed to support data capture for research studies. These children and their parents have been followed for over 13 years, and > 27,000 variables per participant collected and managed in CHILDdb.ca (>90 million data points total), reflecting very diverse characteristics including nutrition, lifestyle, health, environment, mental health, microbiome, metabolome, socioeconomic status and more. During the pandemic, a series of four questionnaires (see Fig. 1) were completed (1: Jan-Jun 2021, 2: Aug-Sep 2021, 3: Oct-Dec 2021, 4: Jan-Mar 2022) by a subset of 1,462 cohort children and their families (5,378 individuals) to assess life changes, stress, coping mechanisms, health, COVID-19 infection rates and attitudes toward vaccines [18].

**Figure 1:**
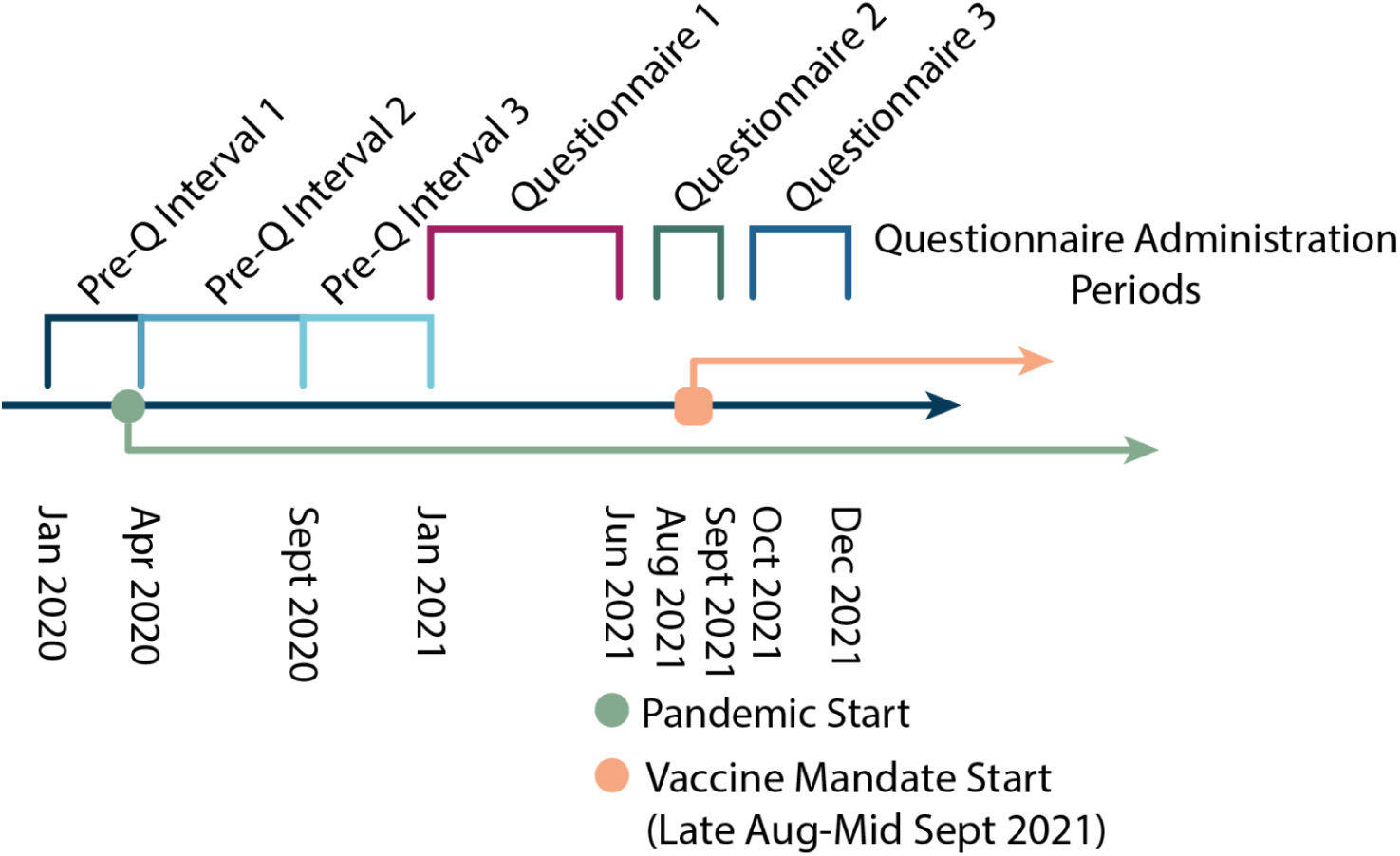
CHILD COVID-19 Add-On Study Timeline. Participants’ responses to three questionnaires were analyzed. The first questionnaire was administered from Jan-Jun 2021, the second was administered from Aug-Sept 2021 and the third from Oct-Dec 2021. During the first questionnaire, participants answered questions about their current beliefs and behaviours, as well as their behaviour during three pre-questionnaire intervals. The first interval was from Jan-Apr 2020, the second was from Apr-Sept 2020 and the third was from Sept 2020 to the time of first questionnaire administration. The start of the pandemic is indicated on the timeline by a green dot, while the start of Canadian vaccine mandates in the regions where CHILD Study participants resided is indicated by a peach square (Late August - Mid September 2021).

### Data Preparation

The COVID-19 add-on study contains participants’ responses to hundreds of questions answered at four time points. (See Fig. 1 for an overview of when questionnaires were administered.) However, there are a limited number of questions where a sufficient number of participants responded to perform a rigorous analysis without imputing data. There were 1,355 adults who responded with their level of agreement to both of the following statements in the 3rd questionnaire (Oct-Dec 2021): “Getting myself vaccinated is important for the health of others in my community” (“community” statement) and “Getting vaccinated is a good way to protect myself from disease” (“self” statement). Participants’ responses to these questions were used as the outcome variables of the study. The third questionnaire was used as the primary outcome as participants’ response rate greatly declined in the fourth questionnaire (Jan-Mar 2022). All data from the fourth questionnaire were omitted from the analysis since the third questionnaire was used as the primary outcome.

Responses to both “community” and “self” questions were re-coded from a 1-4 Likert scale into binary variables for logistic regression modelling. In the original questionnaire, possible responses were “strongly agree”, “somewhat agree”, “somewhat disagree” and “strongly disagree”. These were converted to 1/agree (strongly agree or somewhat agree) and 0/disagree (somewhat disagree or strongly disagree). There was a large class imbalance between those who agreed with the vaccine statements and those who disagreed (∼20:1). Therefore, an effort was made to assemble a set of survey questions to serve as predictor variables where only questions that were answered by all participants who disagreed with at least one vaccine statement were included. This resulted in a dataset of 913 predictor variables (n=131 variables removed). Any individuals who agreed with both vaccine statements and who did not respond to all of these questions were removed from the analysis (n=123 participants removed). This gave a total of 1,232 participants. Although some participants were from the same household, the number of participants per household was not large enough to analyze specific households as a factor.

Questions (variables) were re-encoded where necessary (Likert scales re-ordered, text responses one-hot encoded) using the scikit-learn [30] library. Some questions were removed that were not informative for the purpose of this study (e.g. “which vaccine did you receive?”, since this study is focused on factors that are associated with vaccine confidence or hesitancy in general). Where two variables were >80% correlated, the second variable in the dataset was removed. The data were then scaled using scikit-learn’s standard scaler function. Variables that were not full-rank were removed to eliminate multicollinearity, then variables with variance (s^2^) < 0.16 were removed. This left 686 variables in the dataset (See <u>Supplementary File 2</u>).

scikit-learn’s [30] recursive feature elimination function was then used with the following parameters (estimator = LogisticRegression(class_weight = ‘balanced’), cv = StratifiedKFold(3, shuffle = True, random_state = 5), scoring = ‘f1_macro’) to select the optimal number and set of variables to model responses to each of the following questions: “Getting myself vaccinated is important for the health of others in my community” and “Getting vaccinated is a good way to protect myself from disease’’. The “community” question resulted in a set of 95 variables, while the “self” question resulted in a set of 33 variables.

### Statistical Analyses

In order to determine whether education level, income level, sources of COVID-19-related information, COVID-19 and influenza vaccination status were significantly associated with opinions about vaccination, Fisher exact tests were conducted using the R rstatix package [31] and p-values were adjusted for multiple testing using the Benjamini-Hochberg method.

To determine whether a significant number of individuals had changed their opinions regarding the “community” and “self” statements between the second (Aug-Sept 2021) and third (Oct-Dec 2021) questionnaires, and to determine whether a significant number of individuals had changed their COVID-19 vaccination statuses between the second and third questionnaires, McNemar-Bowker tests were conducted using the R rcompanion package [32].

### Data Modelling: scikit-learn models

scikit-learn [30] penalized logistic regression pipelines were run with grid search to tune hyperparameters for both “community” and “self” datasets, then models were built. The pipeline was run 10 times for each dataset so that an average macro-f1 score could be calculated for each model. F1 is calculated as:

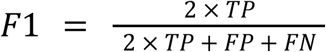

where TP is the number of true positives, FP is the number of false positives and FN is the number of false negatives. Macro-f1 is calculated as:

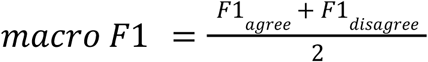

where the f1 scores for those who agree and those who disagree with the statement are calculated separately, then the unweighted mean of the two f1 scores is calculated. This does not take class imbalance into account, and ensures that the score is equally weighted between the two classes (those who agree with the statement and those who disagree). For each iteration of the pipeline, 20% of the data were reserved for model evaluation after hyperparameter optimization, the data were shuffled but stratified by response to the “community” or “self” question, the class weight was balanced and the seed was 6. The following hyperparameters were used for the grid searches of both datasets: {penalty: [‘l1’, ‘l2’, ‘elasticnet’], warm_start: [False, True], solver: [‘liblinear’(l1 penalty only), ‘saga’, ‘lbfgs’(l2 penalty only)], max_iter: [100, 150, 200]}. The same hyperparameters were selected for both models as follows: penalty - l2, solver - lbfgs, max iterations - 100, warm start - false. The grid search pipeline was executed to optimize f1-macro scores of both models. For each model, the proportion of the predictive power of the model that is explained by each domain was calculated as the sum of absolute values of the beta coefficients of each feature belonging to the domain: Σ|β|

These models are referred to in the Results section as the “**scikit-learn models**”.

### Data Modelling: statsmodels models

The absolute values of the beta coefficients from the features of the scikit-learn models above were used as feature weights. The threshold weight for inclusion of a feature in a model was progressively increased by a stepsize of 0.005 in a range of 0.2 to 1.0 such that the number of features in progressive models decreased (see Fig S2). The dataframe containing the feature input values to build each model was the one generated by the “Data Processing” section above. Each model was built using statsmodels [33] with l1 penalization. Odds-ratios, lower and upper 95% confidence intervals, p-values, and false discovery rate corrected p-values were also calculated (using the Benjamini-Hochberg method) for all models. The best models for the “community” and “self” datasets were selected as the model with the most features that had significant corrected p-values (P-adj<= .10) and the fewest total features. These models are referred to in the Results section as the “**statsmodels models**”.

## Results

### Vaccination Beliefs vs Vaccination Actions

This cohort has an average age of 44.1 years (SD = 5.8) and is 62.7% female. 70.9% have a university education (41.8% have a bachelor’s degree and 29.1% have a graduate degree). 44.1% have an annual household income of $150,000 or more. At the CHILD study’s inception, 31.4% were located in Winnipeg, 30.7% were located in Vancouver, 19.5% were in Edmonton and 18.4% in Toronto (See Table S1).

In this cohort, the majority of participants stated that they believed vaccines are important for the health of both their community (“community” statement) (1174, 95.3%) and their own health (“self” statement) (1204; 97.7%) in response to the third questionnaire (administered between October-December 2021, after the implementation of vaccine mandates in some regions), (Fig. 2A). Of these “vaccine-confident” individuals, most who agreed with both the “community” statement (1163 / 1174; 99.1%) and the “self” statement (1180 / 1204; 98.0%) were vaccinated against COVID-19 (Fig. 2B/C). Of the individuals who agreed with the “community” and the “self” statements, 144 / 1174 (12.3%) and 144 / 1204 (12.0%) were vaccinated against influenza, respectively. Roughly two thirds of those who disagreed with the “community” and “self” statements (37 / 58; 63.8%) and (19 / 28; 68.0%) received the COVID-19 vaccine. Only two individuals who disagreed with each statement were vaccinated against influenza (2 / 58; 3.4% and 2 / 28; 7.1%).

**Figure 2:**
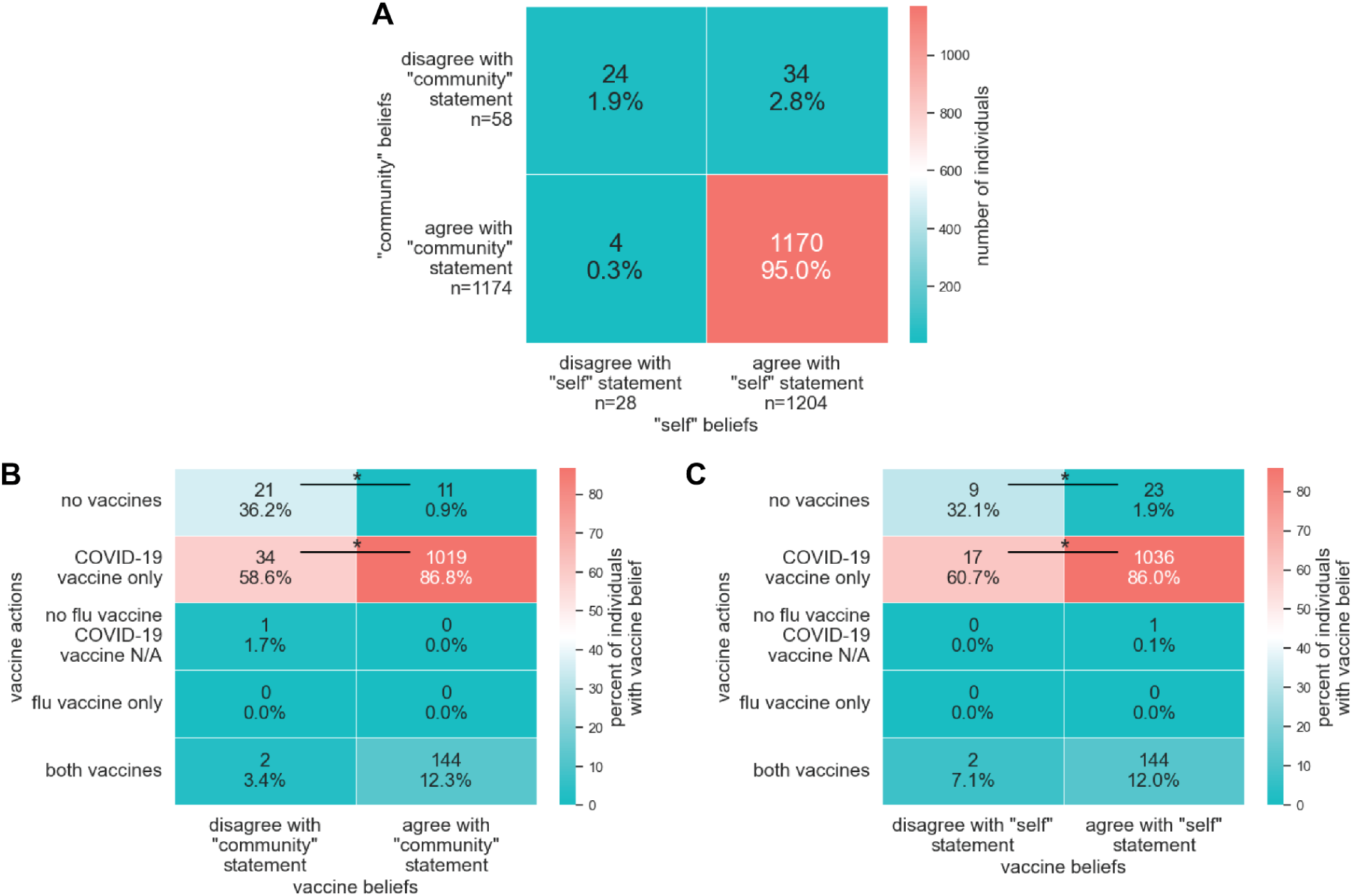
Participants’ vaccination beliefs and vaccination statuses. Participants were asked whether they agreed or disagreed with the following vaccine belief statements: “Getting myself vaccinated is important for the health of others in my community” (“community” statement) and “Getting vaccinated is a good way to protect myself from disease’’ (“self statement”). Participants also responded to the following vaccine action questions: “have you received a dose of a COVID-19 vaccine” and “have you received a dose of influenza vaccine in the last year”. The number of individuals who agreed and disagreed with each vaccine belief statement are shown in panel A. The number of individuals who responded yes/no to the vaccine action questions and agreed/disagreed with the vaccine “community” belief statement are shown in panel B, while the number of individuals who responded yes/no to the vaccine action questions and agreed/disagreed with the vaccine “self” statement are shown in panel C. Significant differences in vaccine actions between participants who have diverging vaccine beliefs (as assessed by a Fisher exact test with correction for multiple testing using the Benjamini-Hochberg method) are denoted by an asterisk (P-adj <= 0.05).

Receiving a COVID-19 vaccination (but not an influenza vaccination as well) was significantly more common among those who agreed with the “community” and the “self” statements than among those who disagreed with them (P-adj = 1.70e-08 and 1.89e-04, respectively) (Fig. 2B/C), and receiving neither the COVID-19 nor the influenza vaccine was more common among those who disagreed with both statements (P-adj = 1.78e-18 and 4.08e-07), as assessed by Fisher exact tests (corrected for multiple testing using the Benjamini-Hochberg method).

The CHILD COVID-19 Add-On study collected data on participants’ agreement with the “community” and “self” statements and COVID-19 vaccination statuses both during (questionnaire 2; Aug-Sept 2021) and after the imposition of COVID-19 vaccine mandates (questionnaire 3; October-December 2021) (mandates were imposed in late-August to mid-September 2021 in the provinces where participants lived [8]). We found that participants’ opinions toward the statements altered over this period (see Fig. 3A/B and Table 2A/B).

**Figure 3A-C:**
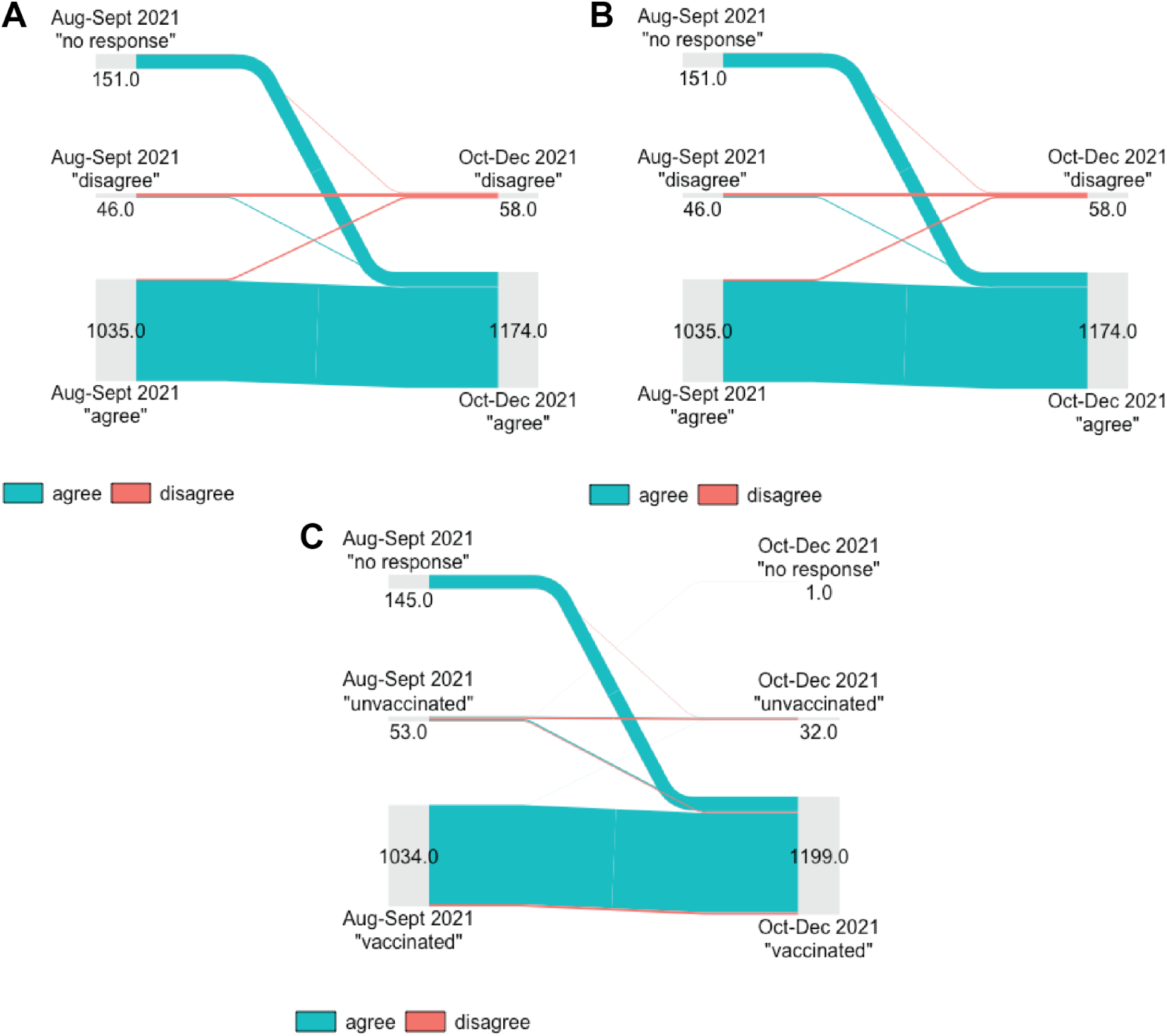
Vaccination opinion statement and COVID-19 vaccination status change between the second and third questionnaires (Aug-Sept 2021) and (Oct-Dec 2021). Participants were asked whether or not they agreed with the following statements: “Getting myself vaccinated is important for the health of others in my community” (“community” statement) and “Getting vaccinated is a good way to protect myself from disease’’ (“self statement”) at two time points: questionnaire 2 (Aug-Sept 2021) and questionnaire 3 (Oct-Dec 2021). Participants’ changes in response to the “community” statement between questionnaire 2 and questionnaire 3 are shown in panel A. Numbers inside or below gray bars indicate the number of participants who had a given response to a questionnaire. Lines connecting grey bars are sized in proportion to the number of participants they represent, and are coloured by participants’ response to questionnaire 3 (Oct-Dec 2021). Participants’ changes in response to the “self” statement between questionnaire 2 and questionnaire 3 are shown in panel B. Participants’ changes in COVID-19 vaccination status between questionnaire 2 and questionnaire 3 are shown in panel C. Lines connecting grey bars are coloured by participants’ agreement/disagreement with the “community” statement in questionnaire 3 (Oct-Dec 2021). For those who changed vaccination status from “unvaccinated” to “vaccinated”, P-adj = 9.63e-06 (proportionally more individuals disagreed with the statement); from “no response” to “unvaccinated”, P-adj = 5.06e-03 (proportionally more individuals disagreed with the statement); from “no response” to “vaccinated”, P-adj = 1.17e-09 (proportionally more individuals agreed with the statement) as determined by via Fisher exact test with correction for multiple testing using the Benjamini-Hochberg method. See **Table S2A/B and S3A-C** for numeric results.

Individuals’ changed significantly toward the “community” statement from “no response” to “agree” (145/151; 96.0%, P-adj = 1.34e-43) and from “no response” to “disagree” (6/151; 1.7%, P-adj = .047). Participants also changed their opinions toward the “self” statement from “no response” to “agree” (148/152; 97.4%, P-adj = 1.68e-144). There were no significant changes from “agree” to “disagree” or “disagree” to “agree” in response to either statement.

Participants who agreed with the “community” statement in questionnaire 3 (1174/1232; 95.3%) had significant changes in vaccination status from “unvaccinated” to “vaccinated” (15/24; 62.5%, P-adj = 7.79e-04), from “no response” to “vaccinated” (139/140; 99.3%, P-adj = 8.61e-42), while those who disagreed with the statement had significant changes in vaccination status from “unvaccinated” to “vaccinated” (10/29; 34.5%, P-adj = 5.85e-03) (see Fig. 3C and Table S3A-C). Among those whose vaccination status changed between questionnaires (173/1232; 14.0%), we assessed whether specific types of status change (e.g. “unvaccinated to vaccinated”) were more common among those who agreed or disagreed with the “community” statement in questionnaire 3. Those who agreed with the statement were more likely to have their vaccination status change from “no response” to “unvaccinated” (139/156; 89.1%, P-adj = 1.17e-09), while those who disagreed with the statement were more likely to have their status change from “unvaccinated” to “vaccinated” (10/16; 62.5%, P-adj = 9.65e-06) and from “no response” to “vaccinated” (3/16; 18.8%, P-adj = 5.06e-03) (see Table S3C).

Our results show that those who disagreed with the vaccine statements were more likely (vs those who agreed) to have their vaccine statuses change to “vaccinated” during the time period when Canadian vaccine mandates were imposed. Given these data and existing literature regarding the efficacy of vaccine mandates in encouraging hesitant individuals to get vaccinated during pandemics (and our supporting data), but failing to ultimately change their beliefs about vaccines ([34] and references therein), and our finding that a significant proportion of individuals’ opinions regarding both the “community” and “self” statements changed between questionnaires 2 and 3, we elected to further examine individuals’ attitudes toward vaccines rather than their vaccination statuses.

### Socioeconomic Status, Education and Sources of Pandemic-Related Information Differ between Vaccine Hesitant and Vaccine Confident Participants

First, we examined whether individuals who agreed and disagreed with the vaccine attitude statements had significantly different sources for COVID-19 related information, levels of formal education or amounts of household income. Among individuals who agreed with the “community” statement (vs. disagreed), a significantly higher proportion completed graduate school (353/1174; 30.1%, P-adj = 3.24e-03), while among those who disagreed (vs. agreed) with the same statement, a significantly higher proportion had only completed high school (12/58; 20.1%, P-adj = 1.34e-03). Among individuals who disagreed with the “self” statement, a significantly higher proportion had completed high school as well (7/28; 25.0%, P-adj = 1.02e-02; Fig. S3).

There were also significant differences in agreement with “vaccines are good for the community” by household income. Those who agreed with the statement were more likely to have a household income above $150,000 (P-adj = 4.72e-05; Fig. S4A). Similarly, there were significant differences in the proportion of individuals who agreed that vaccines are good for oneself based on income. Those who disagreed with the statement were more likely to have a household income between $50,000 - $59,000 (P-adj = 4.39e-02) and those who agreed with the statement were more likely to have a household income above $150,000 (P-adj = 1.69e-02; Fig. S4B).

We also found significant differences in where individuals received information about the pandemic based on their agreement or disagreement with the “community” statement (Fig. S5A). Over half (655/1174, 55.7%) of individuals who agreed with the statement got their information from their provincial government, while only 21/58 (36.2%) of those who disagreed also used this information source (P-adj = 6.7e-03). 14 (24.1%) of individuals who disagreed with the statement got their news from their friends, while only 129 (11.0%) of those who agreed also used this information source (P-adj = 1.58e-02). 20 (34.5%) of individuals who disagreed with the statement got their news from other internet sites (not government websites or social media sites), while only 219 (18.7%) of those who agreed also used this information source (P-adj = 1.29e-02). Those who agreed and disagreed with the “self” statement also differed in where they got their information. 667/1204 (55.4%) of individuals who agreed with the statement got their news from their provincial government, while only 9/28 (32.1%) of those who disagreed also used this information source (P-adj = 2.97e-02). (Fig. S5B). Thus, compared to vaccine-confident individuals, vaccine-hesitant participants were more likely to rely on friends and internet sites not categorized as governmental or social media and less likely to rely on their provincial government for pandemic information.

### Characteristics of Vaccine Hesitant and Vaccine Confident Participants via Logistic Regression

Penalized logistic regression models were built using scikit-learn [30](see Methods) for both the “community” and “self” statement. The scikit-learn models’ macro-f1 scores were 0.95 and 0.93 for the “community” and “self” model, respectively. The “community” model contained 95 features, and the “self” model contained 33 features (See Fig. S6A/B and Table S4A/B). In order to generate models where variables have significant p-values, the number of variables in both models was reduced in a stepwise fashion by increasing the absolute beta coefficient threshold for feature inclusion. Each reduced feature dataset was used to build a penalized logistic regression model in statsmodels (See Methods for description of stepwise model fitting process). The best “community” model contains 19 features of which 12 have odds-ratios >=1.1 or <= 0.9 and corrected p-values <= 0.1. The best “self” model contains 23 features of which 14 have odds-ratios >=1.1 or <= 0.9 and corrected p-values <= 0.1 (Fig. 4A/B). These smaller models were also evaluated for their predictive power using scikit-learn, and have macro-f1 scores of 0.64 and 0.79 for the “community” and “self” model, respectively. A complete list of features, their odd-ratios, upper- and lower-95% confidence intervals, p-values and adjusted p-values from both “community” and “self” models are listed in Tables S5A and S5B, respectively.

**Figure 4:**
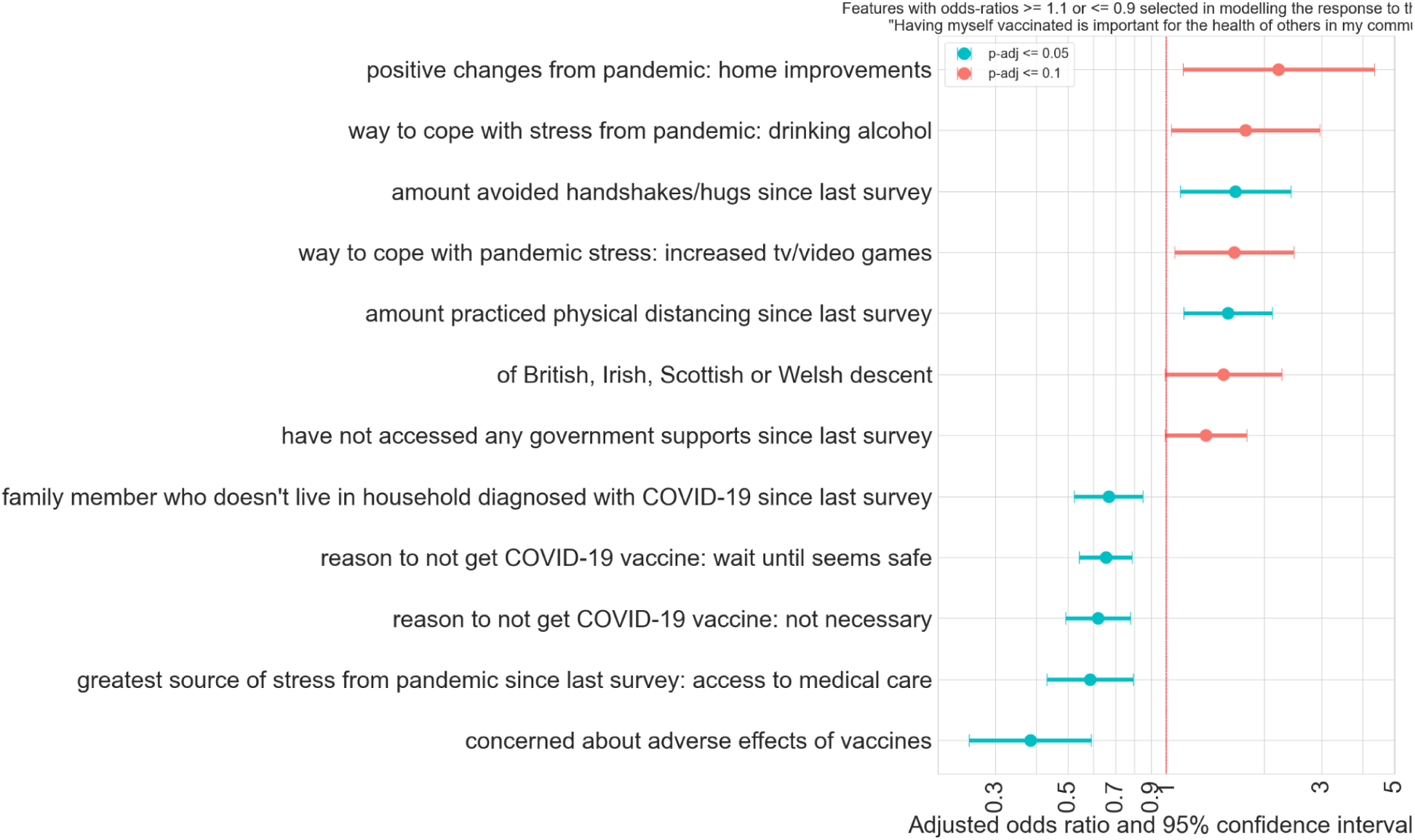
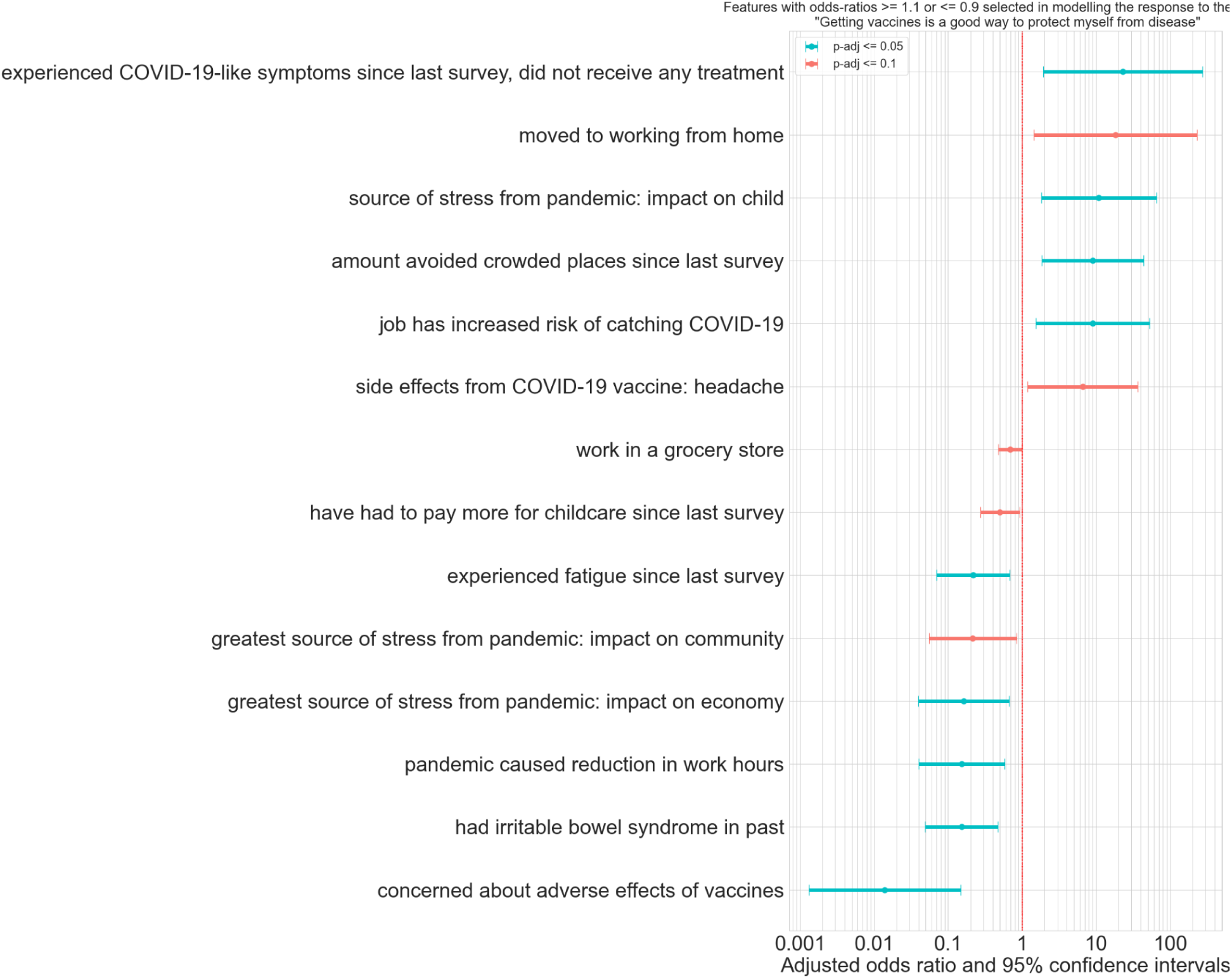
Features from statsmodels logistic regression models agreement & disagreement with the “community” and “self” statements with odds-ratios >= 1.1 or <= 0.9 and significant adjusted p-values. Panels A and B show features from logistic regression models built with participants’ agreement or disagreement with the following statements: “Getting myself vaccinated is important for the health of others in my community” (“community” statement) and “Getting vaccinated is a good way to protect myself from disease’’ (“self statement”) as outcome variables, respectively. Both odds-ratios and 95% confidence intervals are plotted, while significance levels (adjusted p-values) are indicated by the colors (blue : P-adj<=0.05, red : P-adj<=0.10). Features with “amount” in the description are derived from questions that had Likhert scales in the questionnaires.

The results of the scikit-learn models and statsmodels models were concurrent - and the “community” and “self” statement models also overlap. Individuals who agreed with the “community” and “self” statements were both more likely to adhere to COVID-19 safety practices such as physical distancing (OR = 1.55, 95%CI = 1.13 - 2.12, P-adj = .02), have coped with the stress of the pandemic by consuming alcohol (OR = 1.75, 95%CI = 1.04 - 2.96, P-adj = .07), have moved to working from home (OR = 18.23, 95%CI = 1.45 - 229.63, P-adj = .05) and not have accessed government supports (OR = 1.32, 95%CI = 0.99 - 1.77, P-adj = .10). Those who disagreed with the statements were more likely to experience economic precarity (list the pandemic’s impact on the economy as one of their greatest sources of stress (OR = 0.16, 95%CI = 0.04 - 0.67, P-adj = .04) or have had the pandemic cause a reduction in working hours (OR = 0.15, 95%CI = 0.04 - 0.59, P-adj = .04), have experienced or currently experience chronic medical conditions, and have a family member (OR = 0.67, 95%CI = 0.52 - 0.85, P-adj = .01) or have come into contact with an individual who had COVID-19. Those who disagreed with the statements cited concerns about the risk/safety of the COVID-19 vaccine as a reason not to receive the vaccine, and were anxious about the adverse effects of vaccines in general. Many of the participants had front-line occupations. Those whose occupations were historically and relatively well-paid (teachers, health care workers) were more likely to agree with the vaccine statements, while those with occupations that were less well-paid, more precarious and involved working outside the home were more likely to disagree with the vaccine statements (factory workers, grocery store workers).

There are also areas where the “community” and “self” models differ. The “self” model was more focused on highlighting sources of stress experienced by vaccine confident and vaccine hesitant individuals (confident individuals were concerned about the impact of the pandemic on their children, while hesitant individuals were concerned about the impact of the pandemic on their community and the economy). The “community” model was more focused on mechanisms of coping with stress (confident individuals coped by consuming alcohol and watching tv or playing videogames) and highlights multiple concerns that hesitant individuals had with both the COVID-19 vaccine and vaccines in general (they didn’t believe the COVID-19 vaccine was necessary, and didn’t feel that its safety had been proven, and they were worried about adverse effects of any vaccine).

## Discussion

This study provided a rare opportunity to delve into individuals’ attitudes about vaccines from the perspective of their own health and the health of their community before and after the imposition of vaccine mandates in Canada. When combined with diverse data (686 variables), including the status of participants’ employment, mental health, and physical health, we were able to derive key insights into how the pandemic was experienced differently by vaccine hesitant and vaccine confident individuals. These differences could be used to design strategies to reduce vaccine hesitancy and support vaccination campaigns.

### Vaccine Mandates: a Double-Edged Sword

All of the regions of Canada in which CHILD study participants reside announced and implemented COVID-19 vaccine mandates in late August or the first half of September of 2021 [8]. This time period overlaps with the administration of Questionnaire 2. Mandates specified that in order to be employed in certain sectors serving the public, individuals would have to receive a vaccine. These mandates also required that individuals over 12 years of age be vaccinated if they wished to access non-essential businesses and public services (e.g., restaurants and bars). Fig. 3 and Table S3A-C show how participants’ vaccination status changed from Questionnaire 2 (when vaccination mandates were being announced and at the beginning of their implementation) to Questionnaire 3 (when vaccination mandates were fully in place; Fig. 1). Those who disagreed with the “community” statement were more likely (vs. those who agreed) to have received the COVID-19 vaccine between Questionnaire 2 and 3 (P-adj = 9.63e-06), around the time of mandate imposition. Although roughly 60% of vaccine hesitant individuals were vaccinated against COVID-19, their attitude toward vaccination did not change significantly between questionnaires 2 and 3 (P-adj = 0.36 for “community” statement, P-adj = 1 for “self” statement; Fig. 3, Tables S2A/B). Our logistic regression models show that individuals who disagreed with either one or both of the vaccine statements were much more likely to be concerned about the adverse effects of vaccines than those who agreed with the statements.

Vaccine hesitant individuals were also more likely to have specific concerns about the safety of the COVID-19 vaccine. Together, these data suggest that the vaccine mandates implemented in Canada in the late Summer of 2021 [8] were effective in terms of encouraging individuals to become vaccinated even though they did not necessarily believe it was beneficial and had persisting concerns about vaccine safety. Notably, over 140 participants did not respond to the COVID-19 vaccination status question in Questionnaire 2 and this may be because these individuals were hesitant to report they were unvaccinated. Only one participant failed to respond to this question in the 3rd Questionnaire, after vaccine mandates had been imposed.

Analyses conducted by others have confirmed that vaccine mandates are effective tools to increase the percentage of the population who are vaccinated (effects depend on pre-mandate vaccine uptake and vaccine supply), but may have unintended consequences [8,11,12,34,35]. Aside from the fact that vaccine mandates are intended to influence individuals’ actions rather than their attitudes, and will therefore not necessarily result in increased future vaccine uptake (once mandates are lifted), enforcing vaccination can lead to vaccine aversion in those who were previously willing to be vaccinated by choice [27]. This response was not observed in our study, but data were collected for a relatively short period after mandates were imposed (1-5 months, depending on province of residence and Questionnaire 3 administration date).

Influenza vaccination data was also collected from participants in Questionnaire 3. This gives us some insight into vaccine uptake among both vaccine confident and vaccine hesitant individuals when no mandate is in place, and offers some striking contrasts. Among our study participants, the proportion of individuals who received the flu vaccine was not significantly different between those who agreed and disagreed with the vaccine statements (Figure 2). (Among those who agreed with the “community” and “self” statements, roughly 12% had received a flu vaccine within the last year, comparable to the 3.4% who disagreed with the “community” statement and 7.1% who disagreed with the “self” statement. Influenza vaccination proportions overall were much lower than the general Canadian population aged 18-64 (98.6% of participants were below the age of 65 when they answered questionnaire 3), where 29% and 27% received a flu vaccine in the 2020/2021 and 2021/2022 seasons, respectively [36].) According to a Public Health Agency of Canada (PHAC) report, the top reason for not receiving a flu vaccine was believing that it was not necessary [36]. Individuals who disagreed with the “community” vaccine statement were also more likely to believe that the COVID-19 vaccine was unnecessary (odds-ratio=0.62). Therefore, it is possible that unvaccinated vaccine confident individuals did not see the flu vaccine as useful or necessary. The same PHAC report stated that 43% of adults believe that it is possible to get the flu from the flu vaccine. Together, these findings support the need to better communicate the benefits of specific vaccines (e.g., the influenza vaccine specifically) and vaccines in general to the public, while combating misinformation [37–39].

Singh *et al.* conducted a review of the effectiveness of a variety of strategies that have been employed globally to counter vaccine hesitancy and concluded that no single method was superior, but rather, approaches should employ multiple avenues and be customized for the target population [37].

Vaccine mandates in Canada were an effective tool to ensure that 80% of the population became vaccinated against COVID-19 in order to reduce SARS-CoV-2 transmission [40–43]. On both a national and a global scale, there were situations where mandates met with public backlash [44,27] and there is evidence that mandates failed to change the opinions of vaccine hesitant individuals [45]. Mandates are therefore unlikely to be a viable long-term solution to enhance vaccine uptake. Conversely, in the absence of mandates (as with the influenza vaccine), even vaccine confident individuals can struggle to see the benefits of receiving routine immunizations. In order to improve both specific and general vaccine uptake, there exists a need for enhanced public messaging that highlights benefits while dispelling common myths.

### Where You Get Your News Matters: Sources of Pandemic-Related Information and Hesitancy

Our study found notable differences in use of information sources for those who were more vaccine hesitant. Work by Charron *et al.* and Carrieri *et al.* [46,47] has also shown that vaccine hesitant individuals were more likely to rely on both the internet and social media to gain information, as well as depending on their family. They were less likely to rely on healthcare professionals. Our study was more granular in the examination of the use of information sources for COVID-19 related information, as the CHILD questionnaires divided “internet” into “federal government websites”, “provincial government websites”, “social media” and “other internet” (Fig. S5A/B). CHILD questionnaires also provided many more non-internet options for information sources. We found that those who disagreed with the “community” statement were significantly more likely to rely on friends and “other internet” sites for advice, and less likely to rely on provincial government websites than those who agreed with the statement (see Fig S5A/B). Those who agreed with the “self” statement were also significantly more likely to rely on provincial government websites than those who did not. Note that in Canada, healthcare and public health is a provincial/territorial jurisdiction, so this is consistent with seeking information from an authoritative source. Our modelling of responses to the “community” statement also found that those who disagreed with the statement were much more likely to have found access to medical care to be a large source of stress during the pandemic (odds-ratio=0.72). These individuals may not have been able to request personalized vaccination information from their primary care provider or have their vaccine-related questions answered due to this lack of access. Healthcare providers are generally trusted sources of information [48], and severing this connection between patient and provider may have led to incorrect assumptions based on false information. This speaks to the importance of individuals having access to a primary care provider - at a time when such access has become more challenging [49,50]. Also, given that internet usage patterns are changing and misinformation and disinformation is becoming increasingly difficult to detect [51,52], ensuring that credible and readily available vetted information is accessible to individuals with diverse linguistic backgrounds and levels of education is likely pivotal to combatting vaccine hesitancy. Given that individuals who are vaccine hesitant are less likely to visit governmental websites to seek out reliable healthcare information, such information could be shared on sites that hesitant individuals visit more frequently.

### Level of Education & Household Income: Key Correlates of Hesitancy

The large majority of participants (95%) agreed with both the “community” and the “self” statements. As expected, some participants disagreed with either one or both of these statements. However, among those who disagreed with the “community” statement, a larger proportion *agreed* with the “self” statement than disagreed with it (58.6% vs. 41.4%; P-adj = 1.22e-30) (see Fig 2A). This discordance in opinions between benefits for community and self may be due to the fact that herd immunity is a complex and nuanced concept. Studies have shown that roughly one third of Americans lack an understanding of this phenomenon [53,54], and Pfattheicher et. al [15] demonstrated that intention to vaccinate could be increased via informing individuals about herd immunity.

The lower proportion of individuals who were vaccine hesitant in the CHILD cohort (5%) vs the general Canadian population (23%; the proportion of individuals who disagreed with the statement “vaccines are effective” for the Vaccine Confidence Project in 2023 [9]) could be partially explained by several factors. Firstly, the questions are slightly different, and this nuance can affect results. CHILD participants are located primarily in urban areas, and vaccine hesitancy has been disproportionately linked with those living in rural locations [24]. While CHILD participants generally reflect Canadian demographics, like many cohorts a bias exists toward families with more education on average than the general population [19]. They were also willing to engage in a longitudinal scientific study that involves questionnaires, doctor’s visits and biological samples, which perhaps indicates a greater level of trust in the scientific community.

Our findings that those who were vaccine hesitant were likely to have less education and lower income levels align with many previous studies (summarized in [21]). A recent review found that most public vaccine information is tailored to those who read at or above an 8th grade level [55]. The Programme for the International Assessment of Adult Competencies in 2022 found that 49% of Canadians lack reading, writing and comprehension skills required at a high school level [31] and 19% have Level 1 proficiency (the lowest level of proficiency) [57]. Therefore, increased hesitancy among those with less years of formal education may be at least in part due to the fact that most vaccination information campaigns are targeted at those with higher reading comprehension skills. Further design of such campaigns to reach a broader spectrum of audiences could promote understanding, empathy and vaccine uptake. Groups who have experienced racial, cultural and systemic barriers are more likely to be socioeconomically disadvantaged [25,58,59], and also have historical reasons for distrusting the government [60–62] - which was primarily responsible for promoting COVID-19 vaccine uptake during the pandemic. In these communities, community leaders and organizations were essential for establishing vaccine confidence by working as a trusted link between public health and community members and sharing vaccine information [63]. Thus, one’s formal education level does not necessarily equate with their stance on vaccines. The dissemination of vaccine information via culturally appropriate means is essential to ensure that all segments of the population are reached.

### Recommendations for Reducing Vaccine Hesitancy

This study has provided more detailed insight into differences between vaccine hesitant and vaccine confident individuals. These findings can be translated into strategies to decrease hesitancy. More targeted outreach strategies could be formulated to reach individuals with all levels of literacy and with diverse cultural backgrounds. Messaging campaigns could shift away from relying solely upon traditional platforms such as government websites for information dissemination. The way that people engage with media is constantly evolving, and in order to remain relevant in a world where we are constantly bombarded with conflicting, extraneous information, those responsible for ensuring that we are informed about our health options will also have to adapt. It is equally important to ensure that vaccine outreach campaigns targeted toward communities that have experienced historical inequity involve community leaders and organizations in their work in order to establish trust. Ensuring that the critical link between patient and primary care provider is not broken will also serve to convey vital knowledge.

We identified a significant increase in COVID-19 vaccine uptake in vaccine hesitant individuals (vs confident individuals) following the imposition of Canadian vaccine mandates. However, hesitant individuals who were vaccinated did not change their beliefs about vaccines. If individuals’ vaccine hesitancy is not countered, they are unlikely to seek out booster shots, newly available vaccines or vaccinate their children. This study suggests that countering vaccine hesitancy itself is needed, rather than focusing on having individuals receive a vaccine dose as part of a mandate. Directly addressing the concerns leading to vaccine hesitancy, coupled with adequate healthcare access, and messaging suitable for different education levels, could be an effective strategy to increase a population’s vaccination rate long-term.

## Limitations

As has been described previously, this cohort reflects Canadian demographics, except it contains a higher proportion of individuals who hold university degrees [19].

This fact may be inversely correlated with the small proportion of vaccine hesitant individuals in the cohort. We have taken steps to address this in our machine learning, by ensuring that vaccine hesitant and vaccine confident individuals were equally weighted despite differences in actual numbers. The cohort has also self-selected for individuals who are willing to engage in ongoing scientific research, and may thus have a higher level of trust in science. Nevertheless, this study did identify, and was able to characterize, participants that have clearly indicated vaccine hesitancy in the study. This analysis incorporated a significant number of variables, enabling further identification of key insights that can be used to improve vaccine confidence among Canadians.

## Acknowledgements

We thank the CHILD Cohort Study (CHILD) participant families for their dedication and commitment to advancing health research. Visit CHILD at childstudy.ca.

We would like to thank Luisa Mercado Mendoza for thoughtful conversations regarding machine learning analyses.

## Funding Statement

We are grateful to all the families who took part in this study and the whole CHILD team, which includes interviewers, nurses, computer and laboratory technicians, clerical workers, research scientists, volunteers, managers, and receptionists. The CHILD study was initially supported by the Canadian Institutes of Health Research (CIHR), the Allergy, Genes and Environment (AllerGen) Network of Centres of Excellence (NCE), Don & Debbie Morrison.

CHILDdb and the 8-year visit were supported by: Genome Canada, Genome British Columbia, Genome Alberta, BC Children’s Hospital Foundation, the Sick Kids Hospital Foundation, Women and Children’s Health Research Institute (University of Manitoba), Simon Fraser University, Compute Canada, the Ontario Research Fund, the Schroeder Foundation and the Provincial Health Services Authority.

A CIHR COVID-19 Rapid Research Grant supported the COVID-19 data collection with co-funding from Research Manitoba and the COVID Immunity Task Force (CITF).

E.E.G.’s salary was supported by the Canadian COVID-19 Genomics Network (CanCOGeN; Genome Canada grant #E09CMA) and the Coronavirus Variant Rapid Response Network (CoVaRR-Net), supported by Genome Canada, Innovation, Science and Economic Development Canada (ISED) and Canadian Institutes of Health Research (CIHR) (grant #ARR-175622).

F.S.L.B. is an SFU distinguished professor. M.B.A. holds a Tier 2 Canada Research Chair and is a CIFAR Fellow in the Humans and the Microbiome program.

S.E.T. holds a Tier 1 Canada Research Chair in Pediatric Precision Health and the Aubrey J. Tingle Professor of Pediatric Immunology.

Funding for this study came in part from Genome Canada and Genome British Columbia (grant to S.E.T. [274CHI]), BC Children’s Hospital Foundation, the Provincial Health Services Authority, and the Canadian Institutes of Health Research (grants to S.E.T. [OGB-198237 and OGB-185749]).

## Data Availability

Data described in the manuscript are available by registration to the CHILD database https://childstudy.ca/childdb/ and the submission of a formal request. More information about data access for the CHILD Cohort Study can be found at https://childstudy.ca/for-researchers/data-access/. Researchers interested in accessing CHILD Cohort Study data for their research should contact.

## Author Contributions

**Erin E. Gill:** Conceptualization, Methodology, Software, Validation, Formal analysis, Investigation, Resources, Data curation, Writing - original draft, Writing - review & editing, Visualization, Project administration. **Geoff L. Winsor:** Methodology, Software, Validation, Data curation, Writing - review & editing, Visualization. **Baofeng Jia:** Methodology, Software, Validation, Writing - review & editing, Visualization. **Justin Cook:** Resources, Data curation, Writing - review & editing. **Larissa Lotoski:** Validation, Data curation, Writing - review & editing. **Maria V. Medeleanu:** Writing - review & editing. **Erica Di Ruggiero:** Writing - review & editing. **Emily Cameron:** Writing - review & editing. **Vera Dai:** Writing - review & editing. **Charisse Petersen:** Writing - review & editing. **Marc-André Langois:** Writing - review & editing, Funding acquisition. **Theo J. Moraes:** Writing - review & editing, Supervision, Project administration, Funding acquisition. **Elinor Simons:** Writing - review & editing, Supervision, Project administration, Funding acquisition. **Padmaja Subbarao:** Writing - review & editing, Supervision, Project administration, Funding acquisition. **Stuart E. Turvey:** Writing - review & editing, Supervision, Project administration, Funding acquisition. **Meghan B. Azad:** Writing - review & editing, Supervision, Project administration, Funding acquisition. **Fiona S.L. Brinkman:** Conceptualization, Methodology, Validation, Formal analysis, Investigation, Data curation, Writing - original draft, Writing - review & editing, Supervision, Project administration, Funding acquisition.

## Abbreviations

PHAC: Public Health Agency of Canada
WHO: World Health Organization

## Supplementary Material

**Figure S1:**
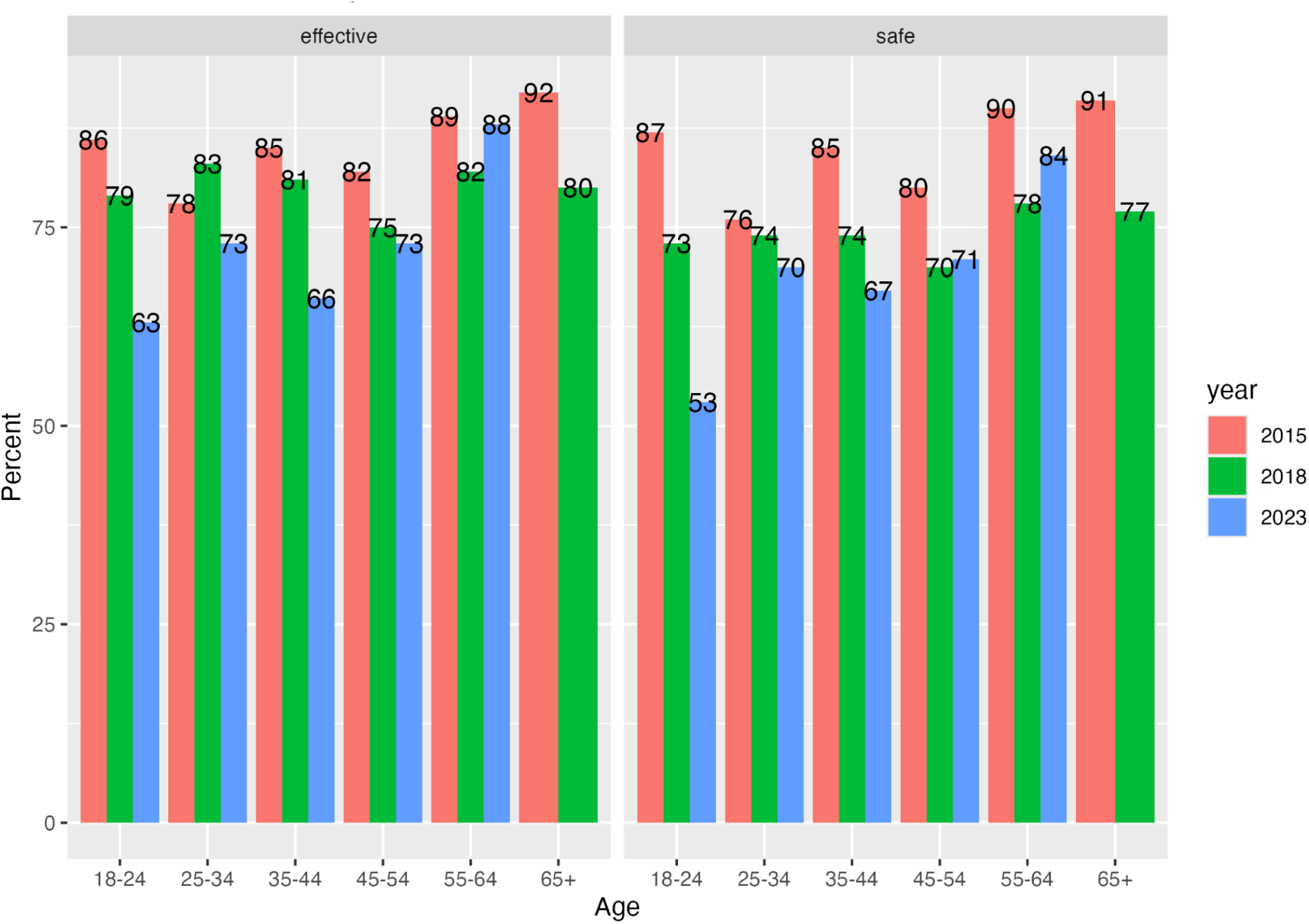
Views on vaccination among Canadians by age before and after the pandemic. The Vaccine Confidence Project (https://www.vaccineconfidence.org) assesses confidence in vaccines in countries around the world. The percentage of Canadian individuals from each age group who agreed with the statement “Vaccines are effective” in three different years is shown on the left, while the percentage of individuals who agreed with the statement “Vaccines are safe” is shown on the right. The data shows a decline in all age groups in confidence in the effectiveness and safety of vaccines between 2015 and 2023. For the 2023 survey, all individuals aged 55 and over were placed in the same age group (i.e. there was no separate 65+ age group). Data adapted from Vaccine Confidence Project website.

**Figure S2:**
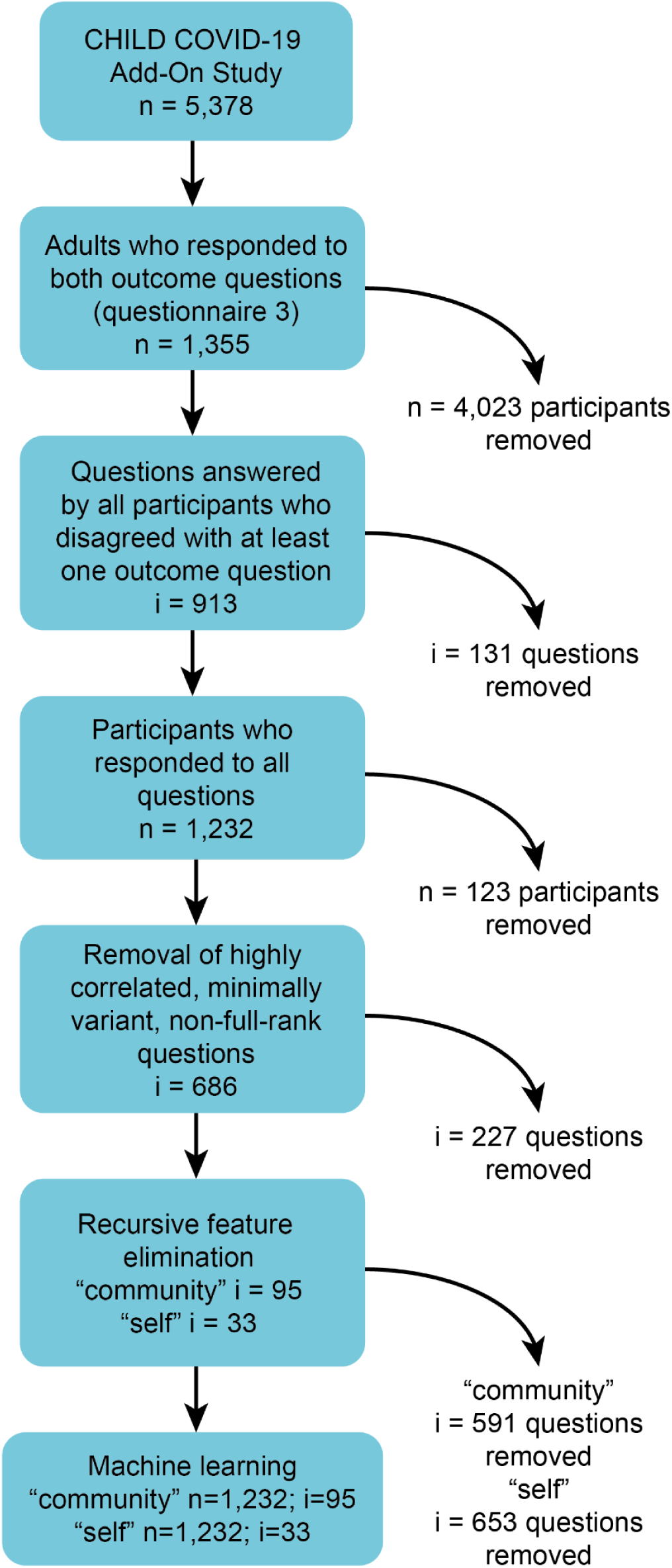
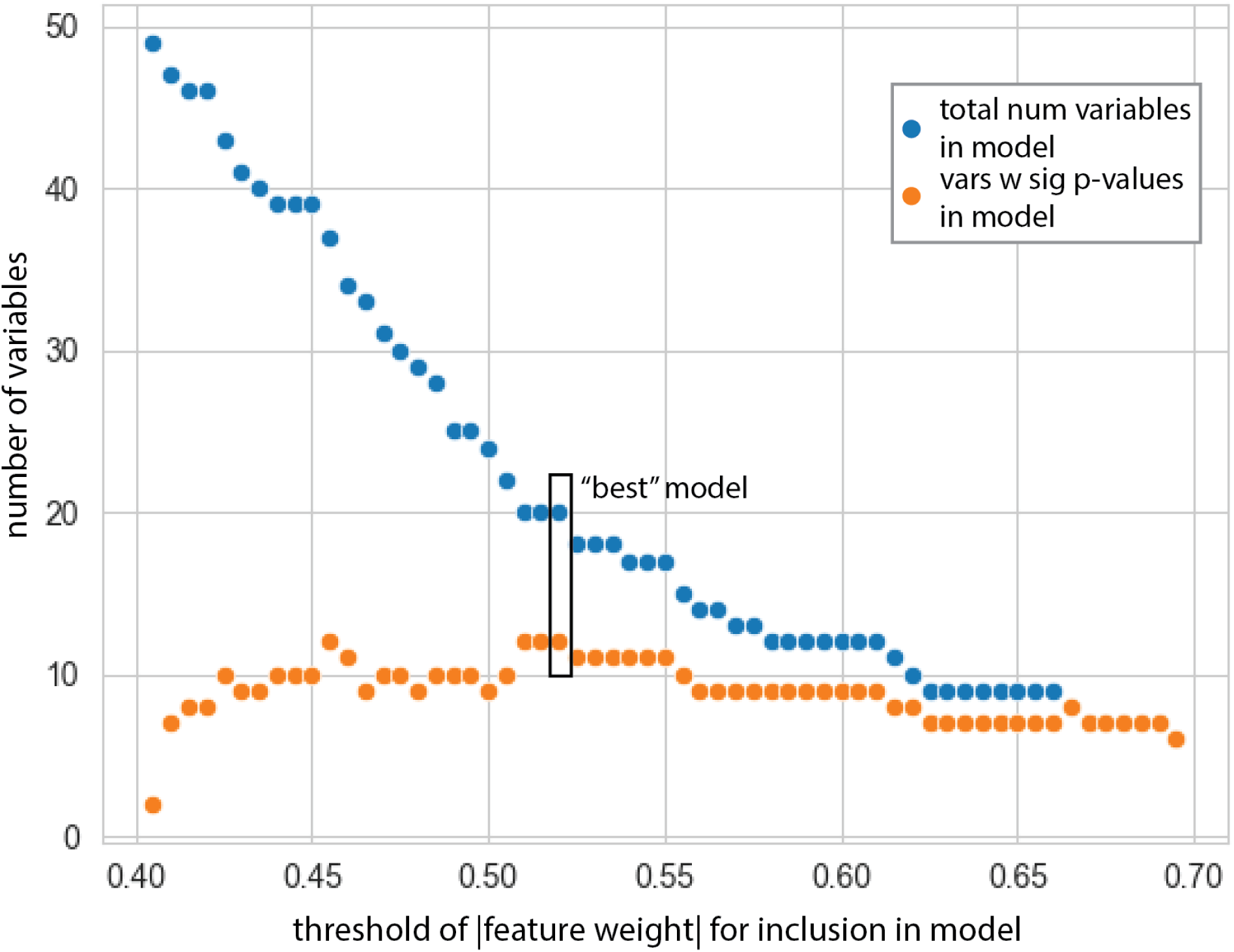
Stepwise approach for fitting the statsmodels models. The absolute values of the beta coefficients from the features of the scikit-learn models were used as feature weights. The threshold weight for inclusion of a feature in a model was progressively increased by a stepsize of 0.005 in a range of 0.2 to 1.0 such that the number of features in progressive models decreased. The total number of features in each successive “community” model are plotted as blue dots, while the number of features in each “community” model that have significant p-values are plotted as orange dots. The best models for the “community” and “self” datasets were selected as the model with the most features that had significant corrected p-values (P-adj <= 0.10) and the fewest total features.

**Figure S3:**
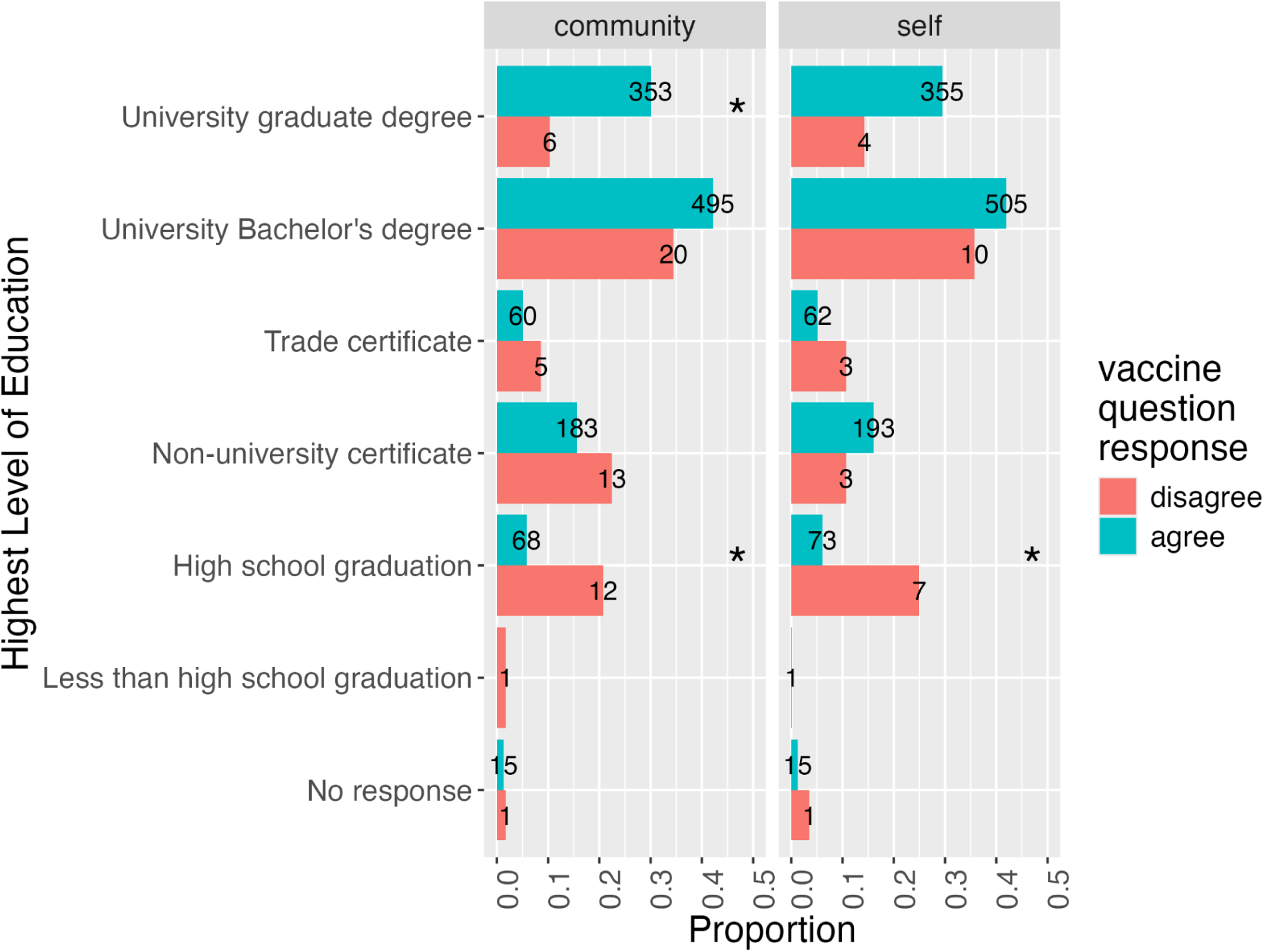
Education level and vaccine hesitancy. Participants’ highest level of completed formal education is grouped by whether or not they agreed with the following statements: “Getting myself vaccinated is important for the health of others in my community” (“community” statement) and “Getting vaccinated is a good way to protect myself from disease’’ (“self statement”). Significance in differences in education levels between those who agreed and disagreed with vaccination statements were determined via Fisher exact test with correction for multiple testing using the Benjamini-Hochberg method. Significant differences (P-adj <= 0.05) between groups of individuals who responded yes and no to the vaccine question are denoted by an asterisk.

**Figure S4:**
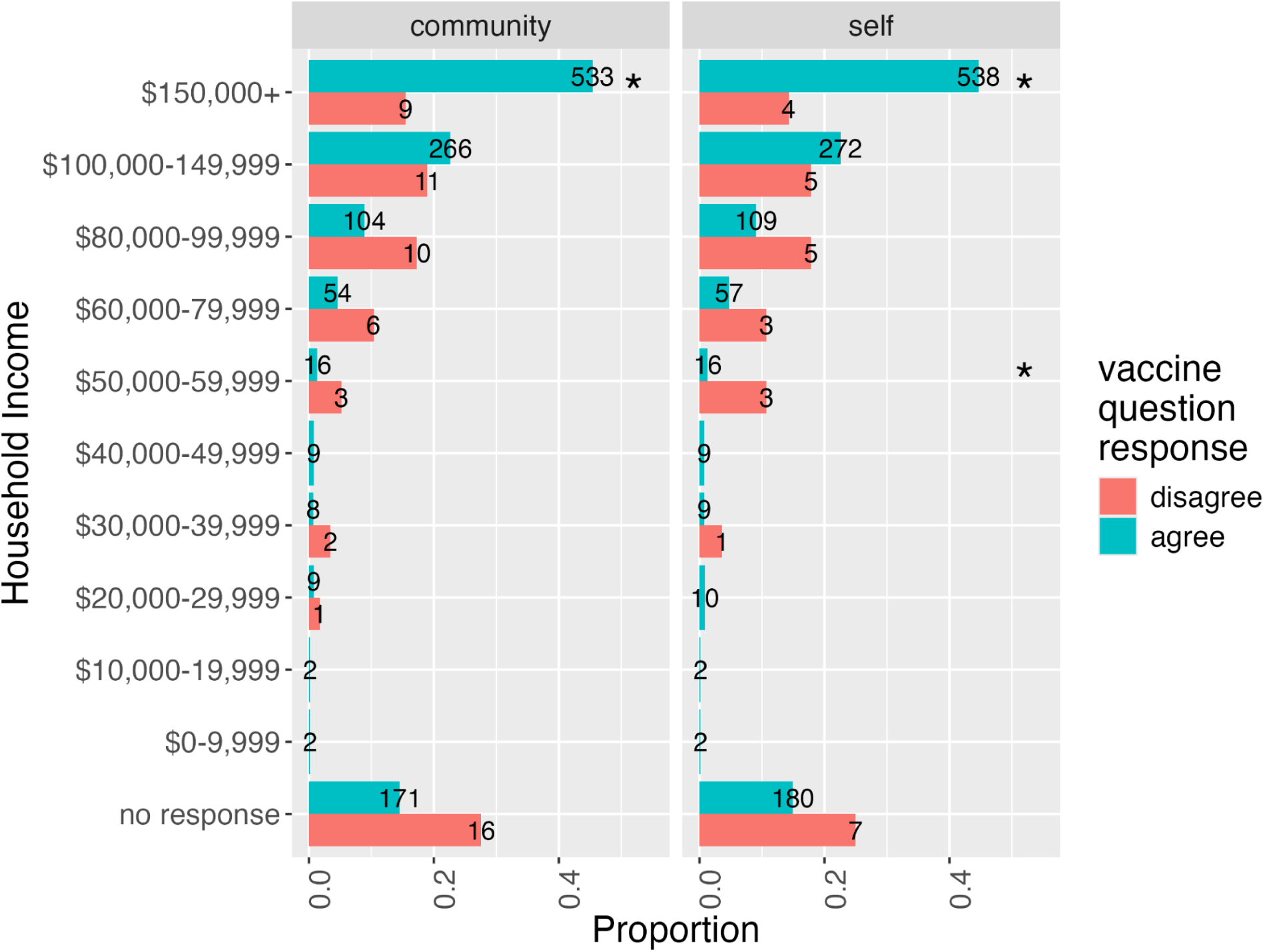
Combined household income and vaccine hesitancy. Participants’ combined household income is grouped by whether or not participants agreed or disagreed with the following statements: “Getting myself vaccinated is important for the health of others in my community” (“community” statement) and “Getting vaccinated is a good way to protect myself from disease’’ (“self statement”) in panels A and B, respectively. Differences between individuals who agreed and disagreed with both statements was determined by Fisher exact test with correction for multiple testing using the Benjamini-Hochberg method. Significant differences (P-adj <= 0.05) between groups of individuals who responded yes and no to the vaccine question are denoted by an asterisk.

**Figure S5A/B:**
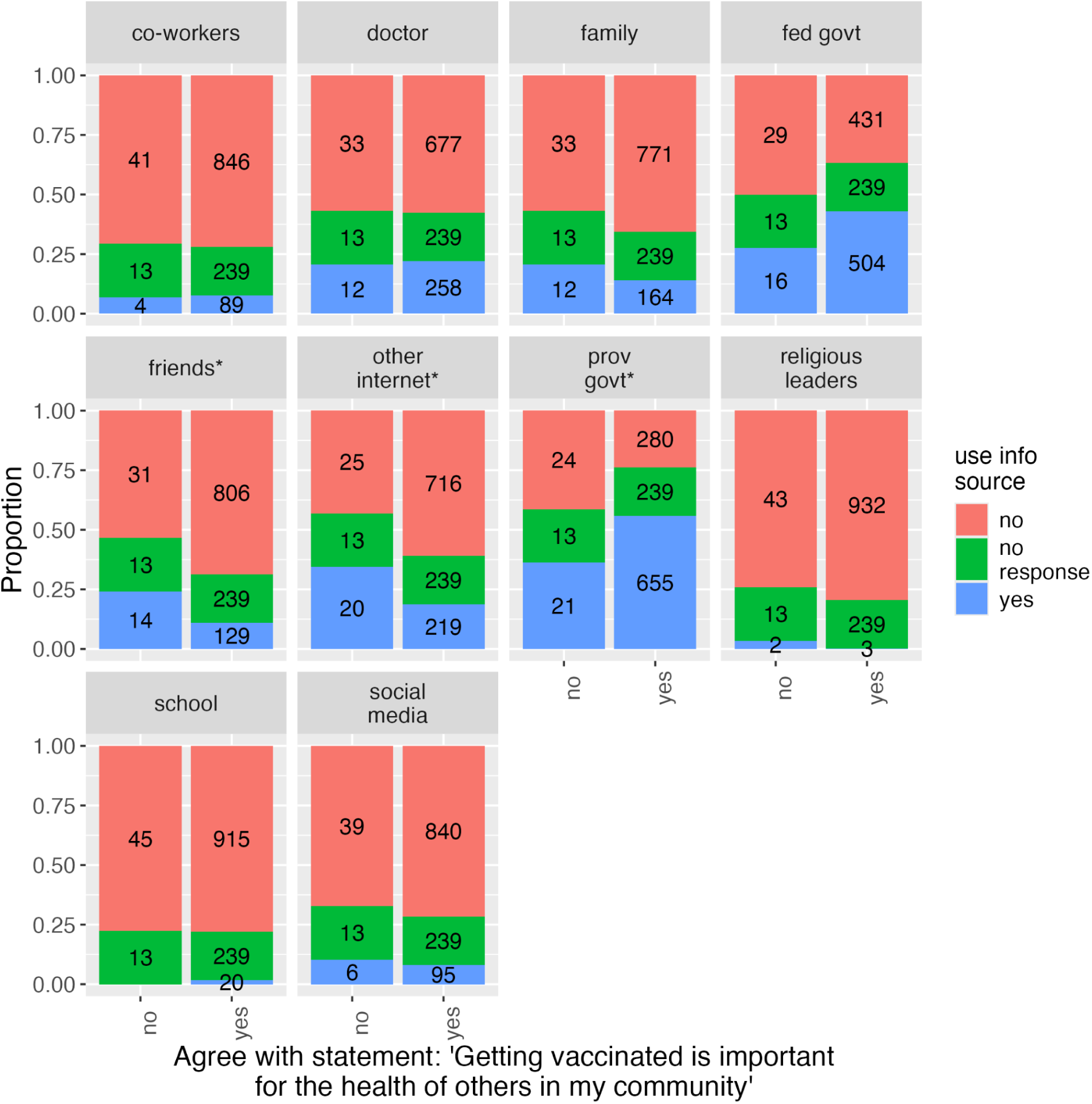
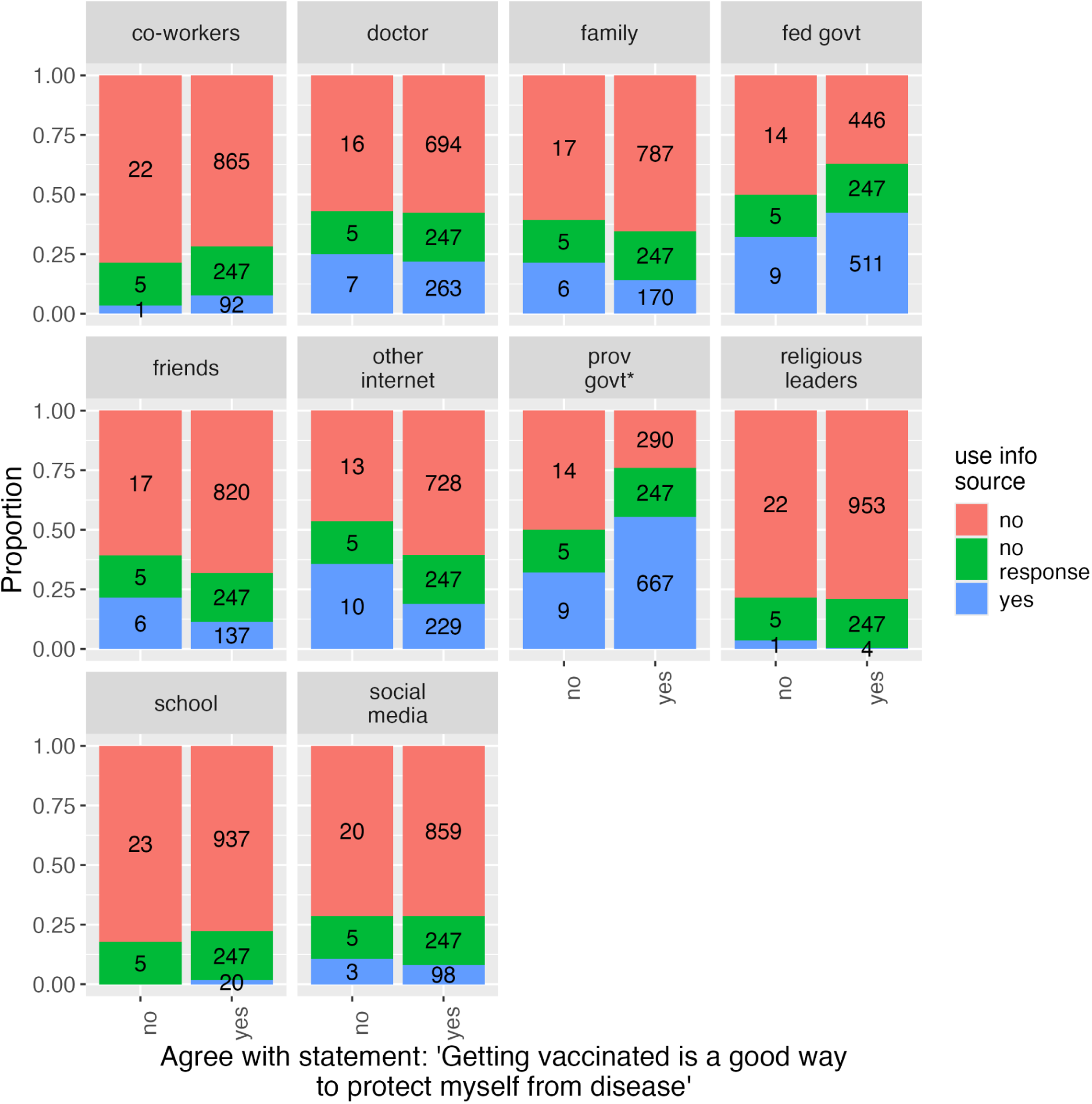
Vaccination beliefs and sources of pandemic-related information. Participants were asked whether they used a variety of sources for information regarding the COVID-19 pandemic. In panels A and B, participants were asked whether they agreed or disagreed with the following statements: “Getting myself vaccinated is important for the health of others in my community” (“community” statement) and “Getting vaccinated is a good way to protect myself from disease’’ (“self statement”), respectively. Proportions of individuals who used/didn’t use each information source are plotted, while actual numbers of individuals are printed on the bars. *Significant difference in usage of news source between individuals who agreed and disagreed with the vaccine statement (Fisher’s exact test with adjusted p-values <= 0.05).

**Figure S6A/B:**
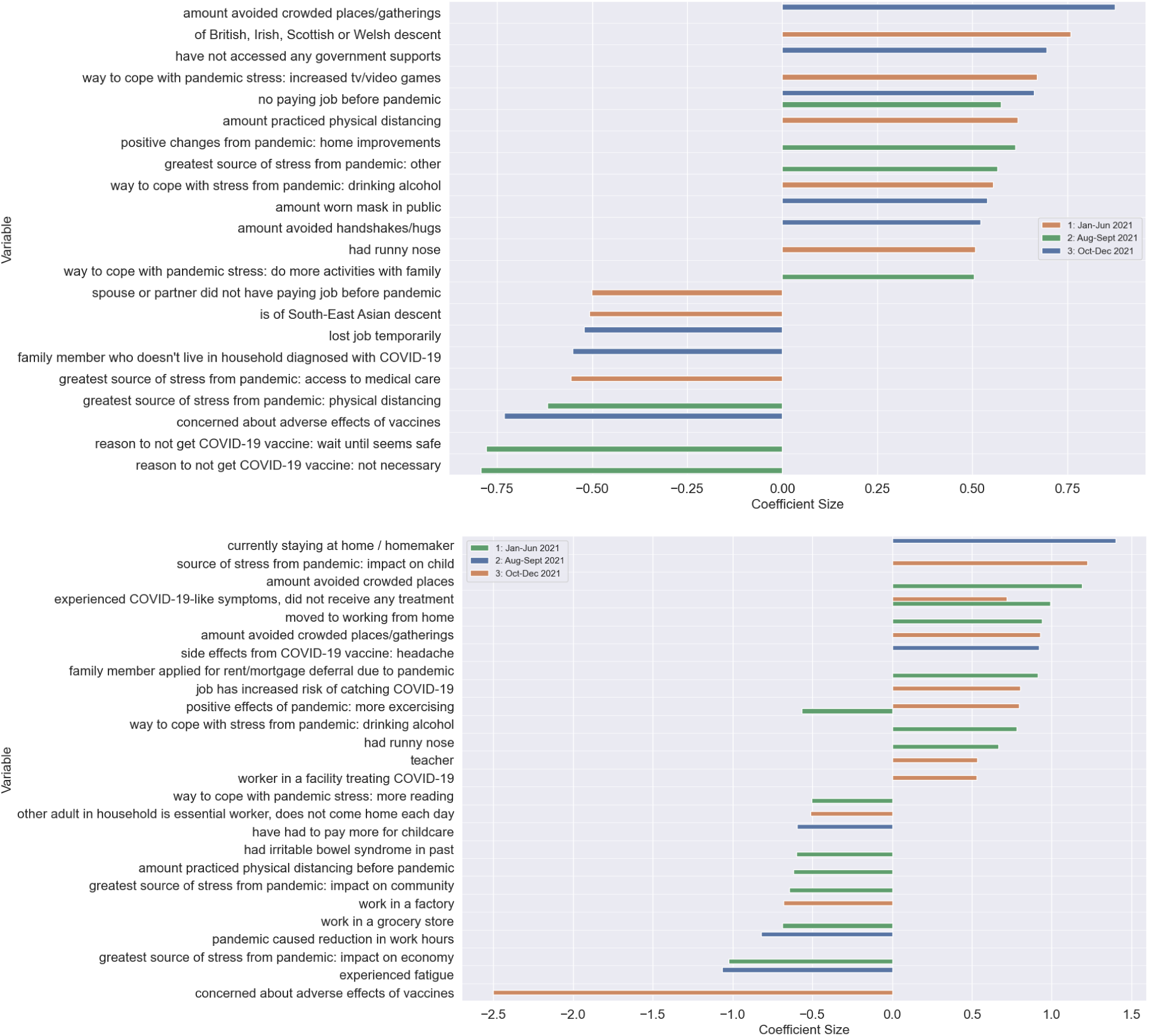
Features from scikit-learn logistic regression models predicting agreement & disagreement with the “community” and “self” statements. Panels A and B show features from penalized logistic regression models built with the outcome variables “Having myself vaccinated is important for the health of others in my community” and “Getting vaccines is a good way to protect myself from disease”, respectively. Colours correspond to the questionnaire from which the feature was derived. Features with absolute coefficients >=0.5 are plotted.

**Table S1:** Sociodemographic characteristics of the cohort (n = 1232)

| Biological Sex at Birth |  |  |
| --- | --- | --- |
| Sex at birth | Number | Proportion |
| female | 773 | 0.63 |
| male | 451 | 0.37 |
| prefer not to answer | 7 | 0.01 |
| prefer to self-describe | 1 | 0.00 |

| Highest Level of Education |  |  |
| --- | --- | --- |
| Highest level of education completed | Number | Proportion |
| less than high school | 1 | 0.00 |
| high school graduation | 80 | 0.06 |
| non-university certificate | 196 | 0.16 |
| trade certificate | 65 | 0.05 |
| bachelor's degree | 515 | 0.42 |
| graduate degree | 359 | 0.29 |
| no response | 16 | 0.01 |

| Annual Household Income |  |  |
| --- | --- | --- |
| Annual household income | Number | Proportion |
| \$0-9,999 | 2 | 0.00 |
| \$10,000-19,999 | 2 | 0.00 |
| \$20,000-29,999 | 10 | 0.01 |
| \$30,000-39,999 | 10 | 0.01 |
| \$40,000-49,999 | 9 | 0.01 |
| \$50,000-59,999 | 19 | 0.02 |
| \$60,000-79,999 | 60 | 0.05 |
| \$80,000-99,999 | 114 | 0.09 |
| \$100,000-149,999 | 277 | 0.22 |
| \$150,000+ | 542 | 0.44 |
| no response | 187 | 0.15 |

| Age |  |  |
| --- | --- | --- |
| Age | Number | Proportion |
| 15-19 | 2 | 0.00 |
| 20-24 | 3 | 0.00 |
| 25-29 | 0 | 0.00 |
| 30-34 | 22 | 0.02 |
| 35-39 | 174 | 0.14 |
| 40-44 | 518 | 0.42 |
| 45-49 | 366 | 0.30 |
| 50-54 | 110 | 0.09 |
| 55-59 | 15 | 0.01 |
| 60-64 | 6 | 0.00 |
| 65-69 | 7 | 0.01 |
| 70-74 | 6 | 0.00 |
| 75-79 | 3 | 0.00 |
|  | mean | 44.06 |
|  | SD | 5.78 |

| Location at Study Inception |  |  |
| --- | --- | --- |
| Location | Number | Proportion |
| Vancouver | 378 | 0.31 |
| Edmonton | 240 | 0.19 |
| Winnipeg | 387 | 0.31 |
| Toronto | 227 | 0.18 |

**Table S2A:** Agreement and Disagreement with the “community” statement in response to questionnaire 2 (Aug-Sept 2021) and questionnaire 3 (Oct-Dec 2021). Table cells where individuals changed their opinion between questionnaires are highlighted in blue. Proportions of individuals whose opinion changed from one response to another between questionnaires are shown in brackets. McNemar-Bowker tests for marginal homogeneity were used to assess whether opinion changes from questionnaire 2 to questionnaire 3 were significant. Significant adjusted p-values (<=0.05) are shown in bold.

| n = 1232 |  | Response to Questionnaire 3 (Oct-Dec 2021) |  |  |
| --- | --- | --- | --- | --- |
|  |  | Disagree<br>n = 58 | Agree<br>n = 1174 | No Response<br>n = 0 |
| Response to<br>Questionnaire 2<br>(Aug-Sept 2021) | Disagree<br>n = 46 | 34 | 12 (26.09%)<br>P-adj = .36 | 0 (0.00%)<br>P-adj = **.047** |
|  | Agree<br>n = 1035 | 18 (1.74%)<br>P-adj = .36 | 1017 | 0 (0.00%)<br>P-adj = **1.34e-43** |
|  | **No Response**<br>n = 151 | 6 (3.97%)<br>P-adj = **.047** | 145 (96.03%)<br>P-adj = **1.34e-43** | 0 |

**Table S2B:**
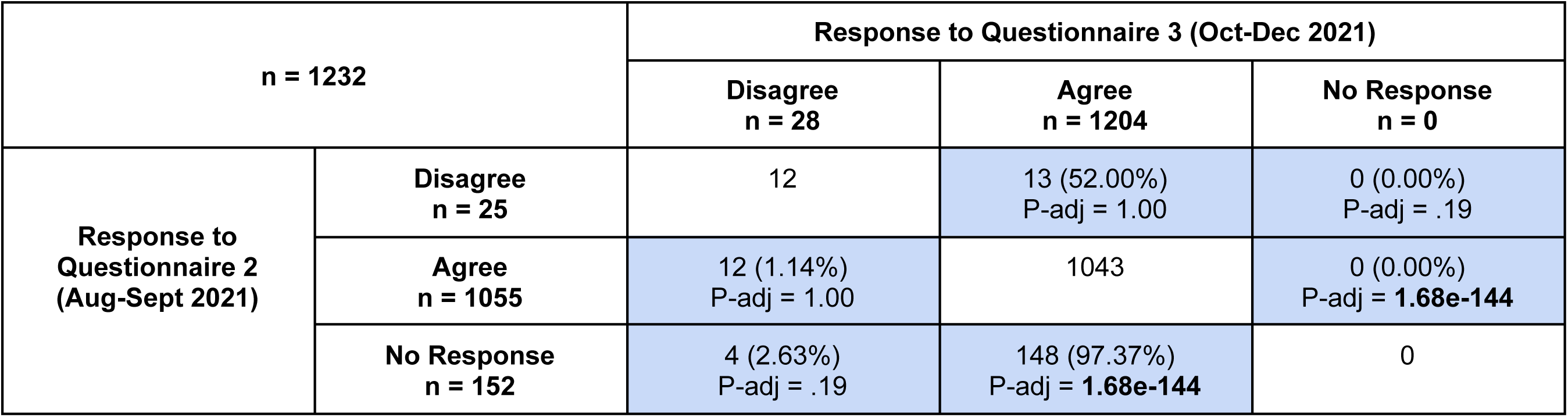
Agreement and Disagreement with the “self” statement in response to questionnaire 2 (Aug-Sept 2021) and questionnaire 3 (Oct-Dec 2021). Table cells where individuals changed their opinion between questionnaires are highlighted in blue. Proportions of individuals whose opinion changed from one response to another between questionnaires are shown in brackets. McNemar-Bowker tests for marginal homogeneity were used to assess whether opinion changes from questionnaire 2 to questionnaire 3 were significant. Significant adjusted p-values (<=0.05) are shown in bold.

| n = 1232 |  | Response to Questionnaire 3 (Oct-Dec 2021) |  |  |
| --- | --- | --- | --- | --- |
|  |  | Disagree<br>n = 28 | Agree<br>n = 1204 | No Response<br>n = 0 |
| Response to<br>Questionnaire 2<br>(Aug-Sept 2021) | Disagree<br>n = 25 | 12 | 13 (52.00%)<br>P-adj = 1.00 | 0 (0.00%)<br>P-adj = .19 |
|  | Agree<br>n = 1055 | 12 (1.14%)<br>P-adj = 1.00 | 1043 | 0 (0.00%)<br>P-adj = **1.68e-144** |
|  | No Response<br>n = 152 | 4 (2.63%)<br>P-adj = .19 | 148 (97.37%)<br>P-adj = **1.68e-144** | 0 |

**Table S3A:** COVID-19 vaccination statuses of participants who agreed with the “community” statement in questionnaire 3 at the time of questionnaire 2 (Aug-Sept 2021) and questionnaire 3 (Oct-Dec 2021). Table cells where individuals changed their statuses between questionnaires are highlighted in blue. Proportions of individuals whose vaccination status changed from one questionnaire to another are shown in brackets. McNemar-Bowker tests for marginal homogeneity were used to assess whether vaccination status changes from questionnaire 2 to questionnaire 3 were significant. Significant adjusted p-values (<=0.05) are shown in bold.

| Response to “community” statement: Agree<br>n = 1174 |  | Response to Questionnaire 3 (Oct-Dec 2021) |  |  |
| --- | --- | --- | --- | --- |
|  |  | Unvaccinated<br>n = 11 | Vaccinated<br>n = 1163 | No Response<br>n = 0 |
| Response to<br>Questionnaire 2<br>(Aug-Sept 2021)<br>n = 1174 | Unvaccinated<br>n = 24 | 9 | 15 (62.5%)<br>P-adj = **7.79e-04** | 0 (0.00%)<br>P-adj = 1.00 |
|  | Vaccinated<br>n = 1010 | 1 (0.10%)<br>P-adj = **7.79e-04** | 1009 | 0 (0.00%)<br>P-adj = **8.61e-42** |
|  | No Response<br>n = 140 | 1 (0.71%)<br>P-adj = 1.00 | 139 (99.29%)<br>P-adj = **8.61e-42** | 0 |

**Table S3B:** COVID-19 vaccination statuses of participants who disagreed with the “community” statement in questionnaire 3 at the time of questionnaire 2 (Aug-Sept 2021) and questionnaire 3 (Oct-Dec 2021). Table cells where individuals changed their statuses between questionnaires are highlighted in blue. Proportions of individuals whose vaccination status changed from one questionnaire to another are shown in brackets. McNemar-Bowker tests for marginal homogeneity were used to assess whether vaccination status changes from questionnaire 2 to questionnaire 3 were significant. Significant adjusted p-values (<=0.05) are shown in bold.

| Response to “community” statement: Disagree<br>n = 58 |  | Response to Questionnaire 3 (Oct-Dec 2021) |  |  |
| --- | --- | --- | --- | --- |
|  |  | Unvaccinated<br>n = 21 | Vaccinated<br>n = 36 | No Response<br>n = 1 |
| Response to<br>Questionnaire 2<br>(Aug-Sept 2021) | Unvaccinated<br>n = 29 | 18 | 10 (34.48%)<br>P-adj = **5.85e-03** | 1 (3.45%)<br>P-adj = .63 |
|  | Vaccinated<br>n = 24 | 0 (0.00%)<br>P-adj = **5.85e-03** | 24 | 0 (0.00%)<br>P-adj = .63 |
|  | **No Response**<br>**n = 5** | 3 (60.00%)<br>P-adj = .63 | 2 (40.00%)<br>P-adj = .63 | 0 |

**Table S3C:**
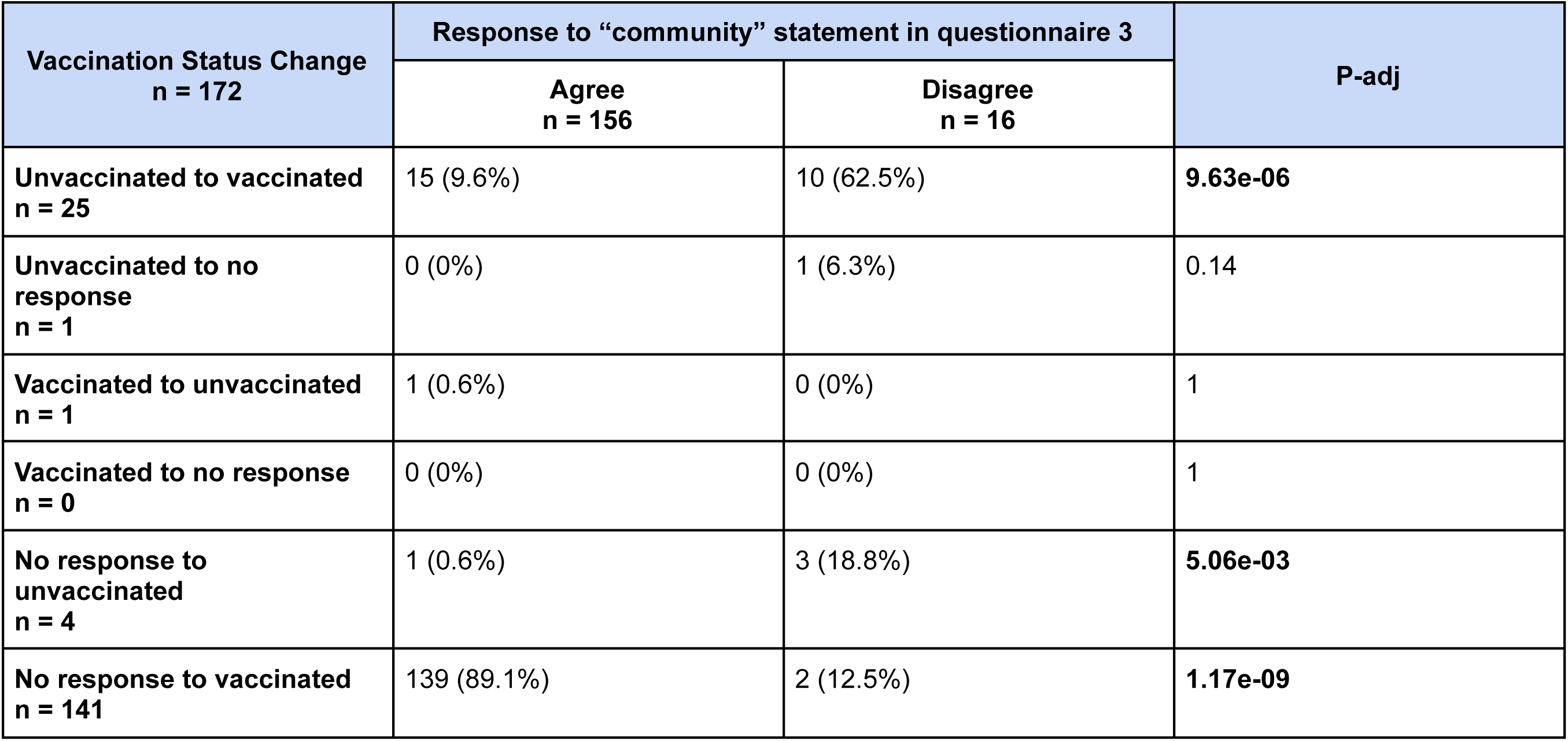
Vaccination status change between questionnaire 2 and questionnaire 3 by response to “community” statement in questionnaire 3. Proportions of individuals who either agreed or disagreed with the “community” statement and had a specific vaccination status change are given in each cell. Differences between individuals who agreed and disagreed with the “community” statement was determined by Fisher exact test with correction for multiple testing using the Benjamini-Hochberg method. Significant differences (P-adj <= 0.05) between groups of individuals who responded yes and no to the vaccine question are shown in bold.

**Table S4A:**
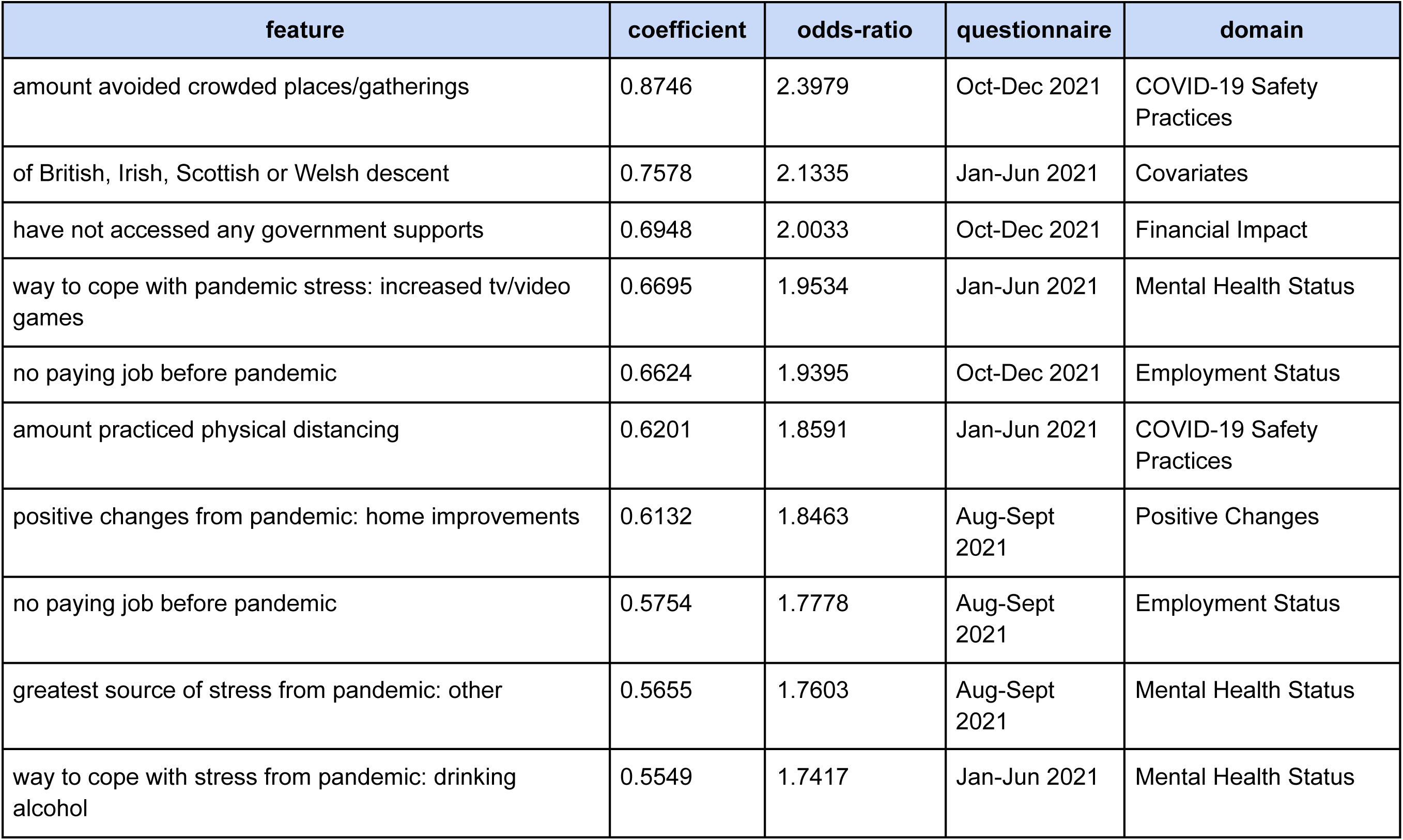
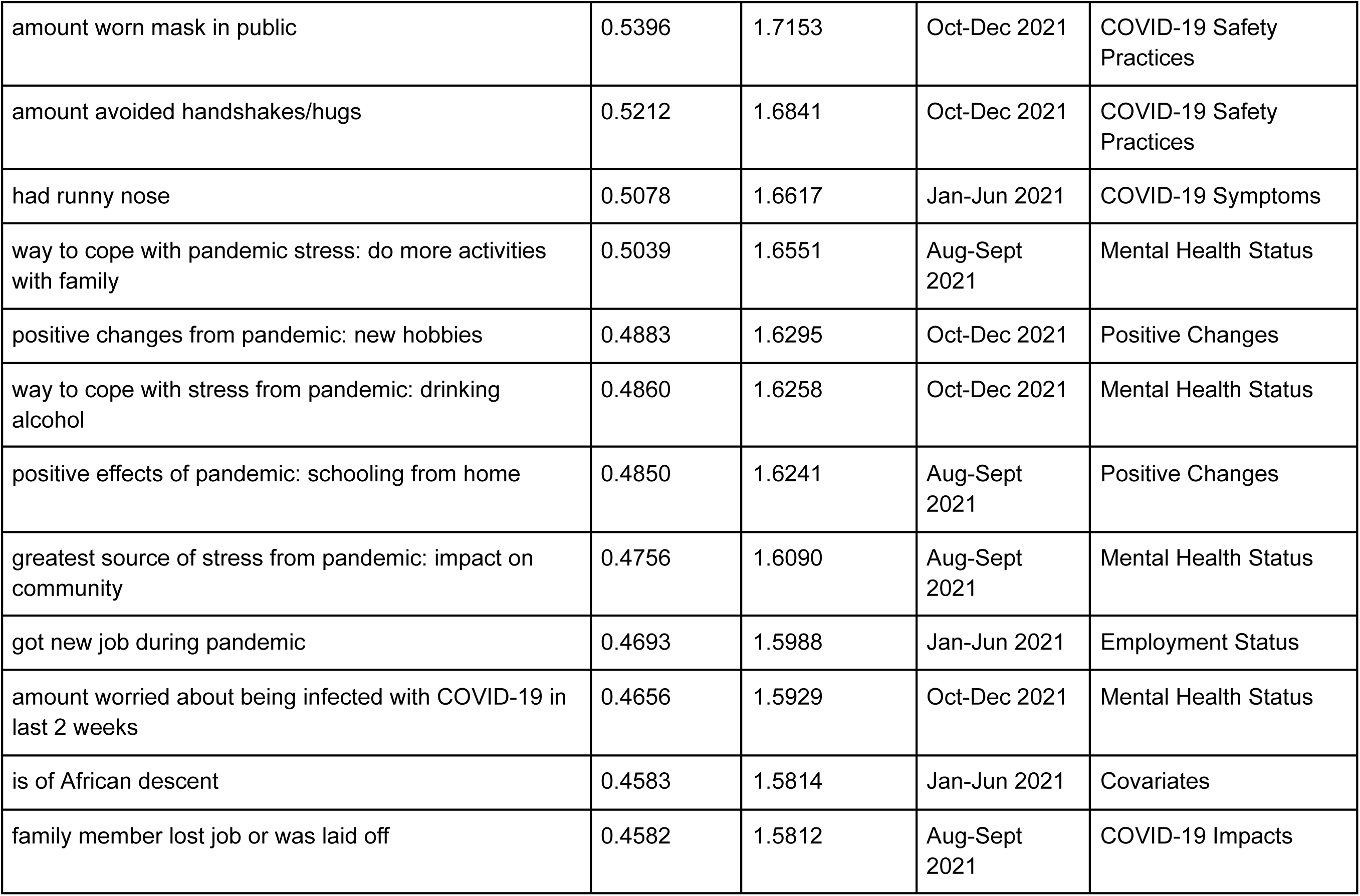
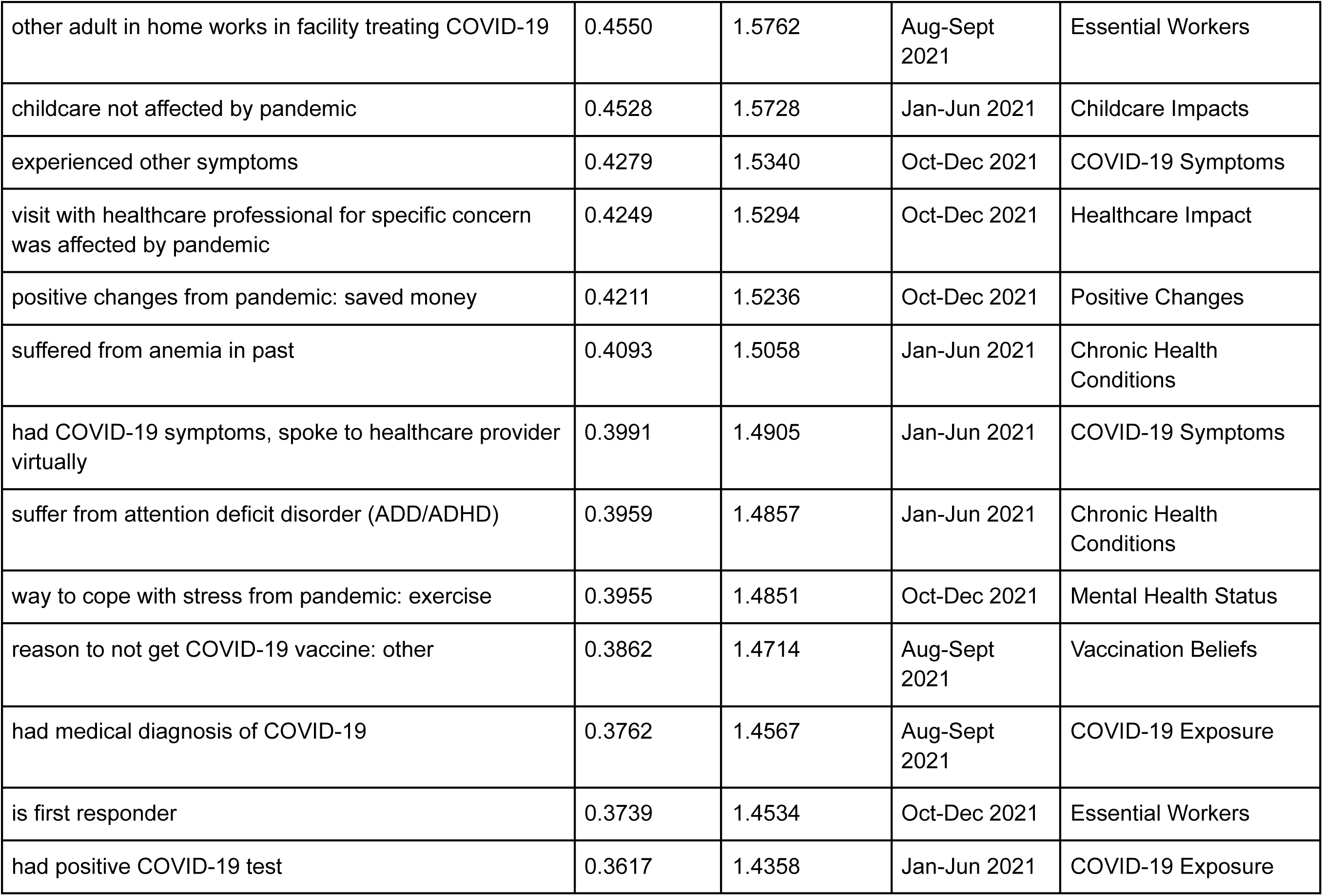
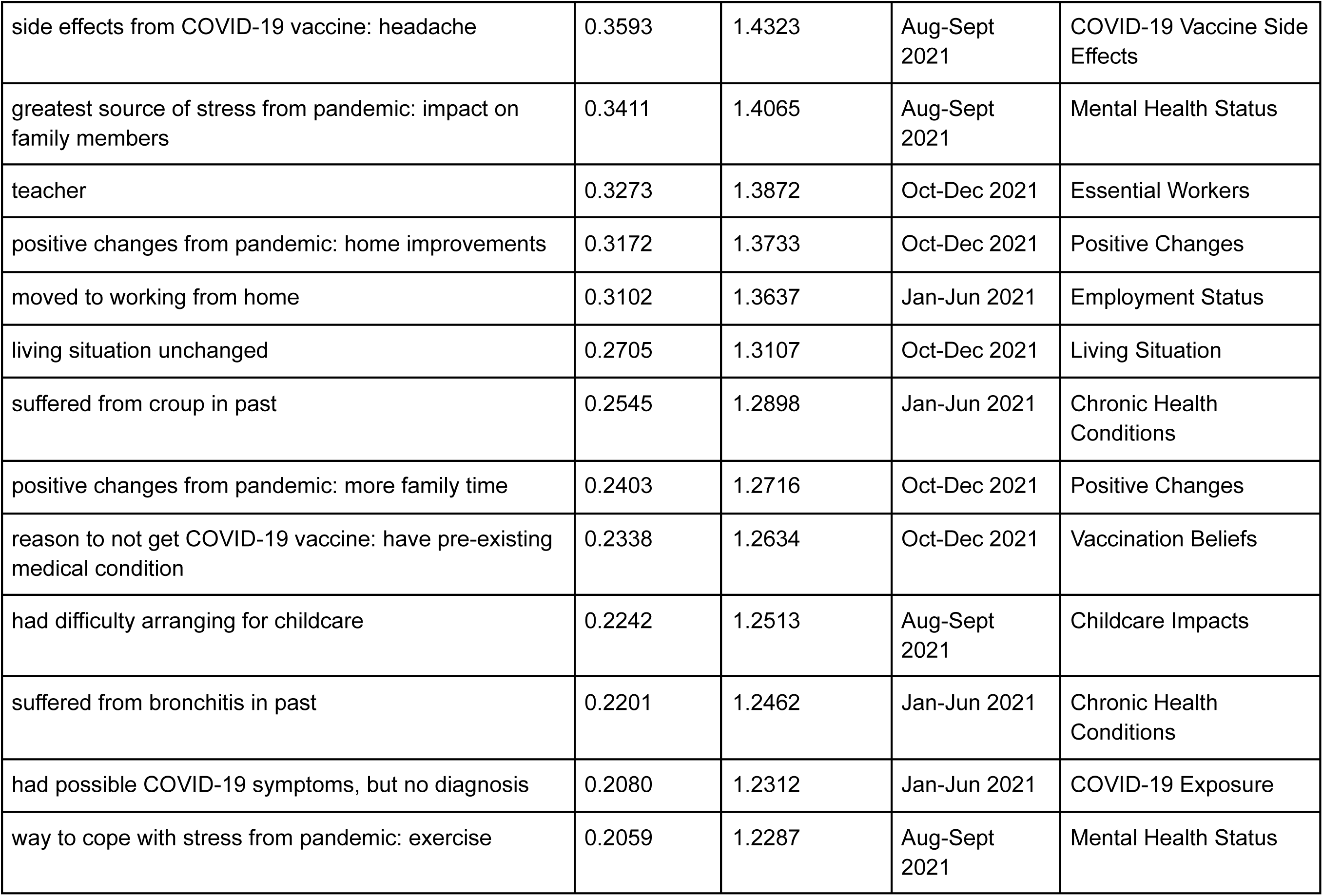
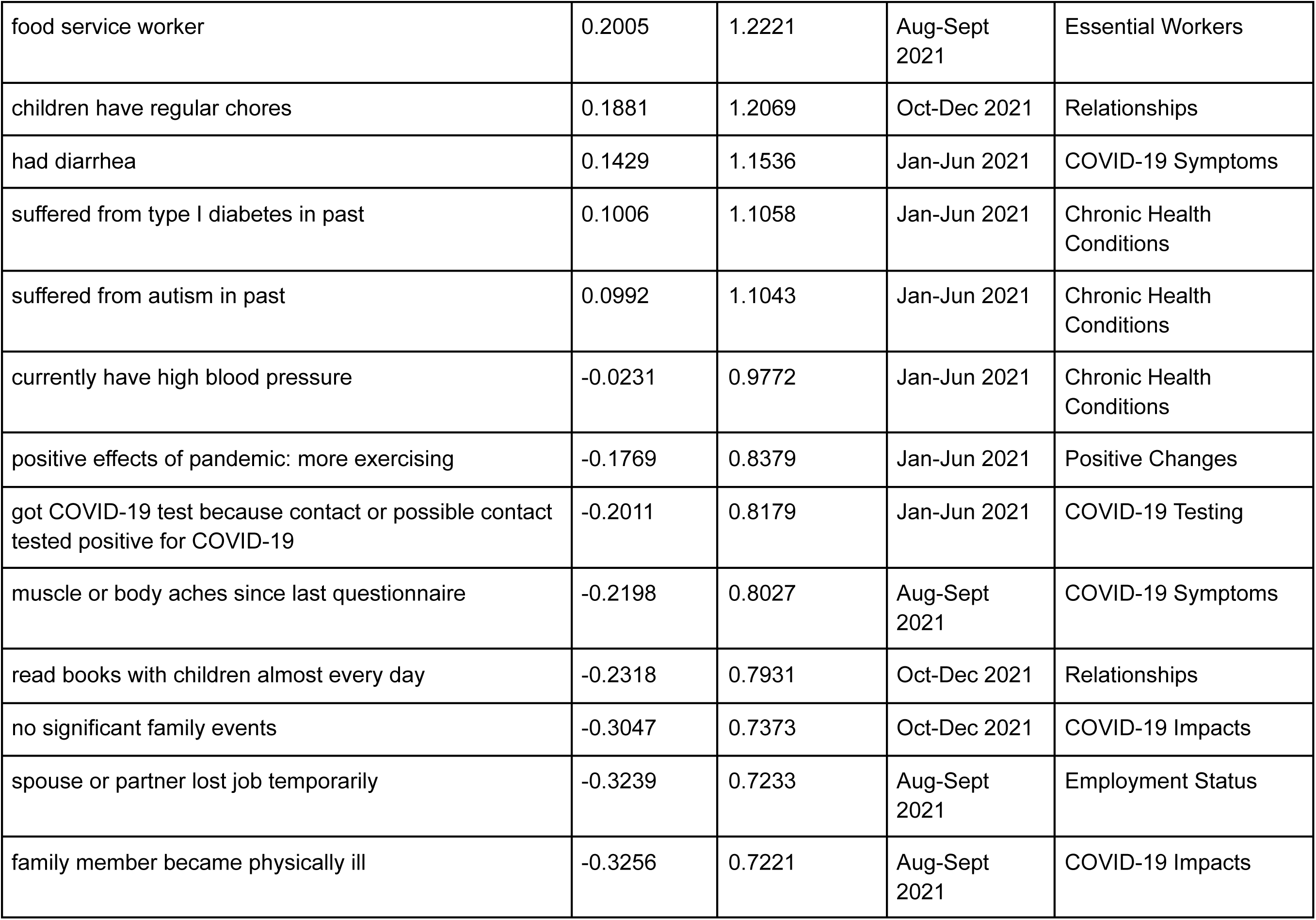
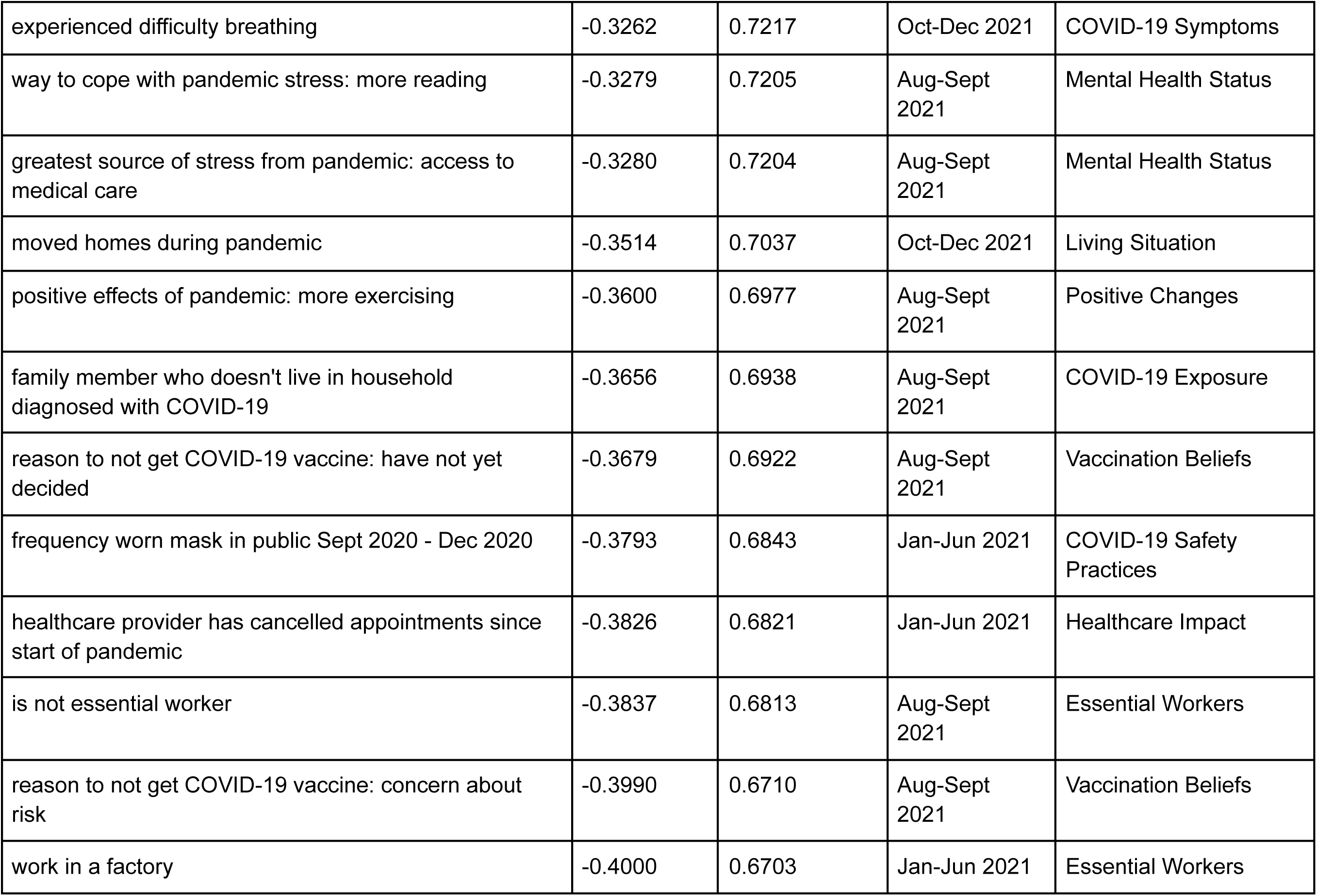
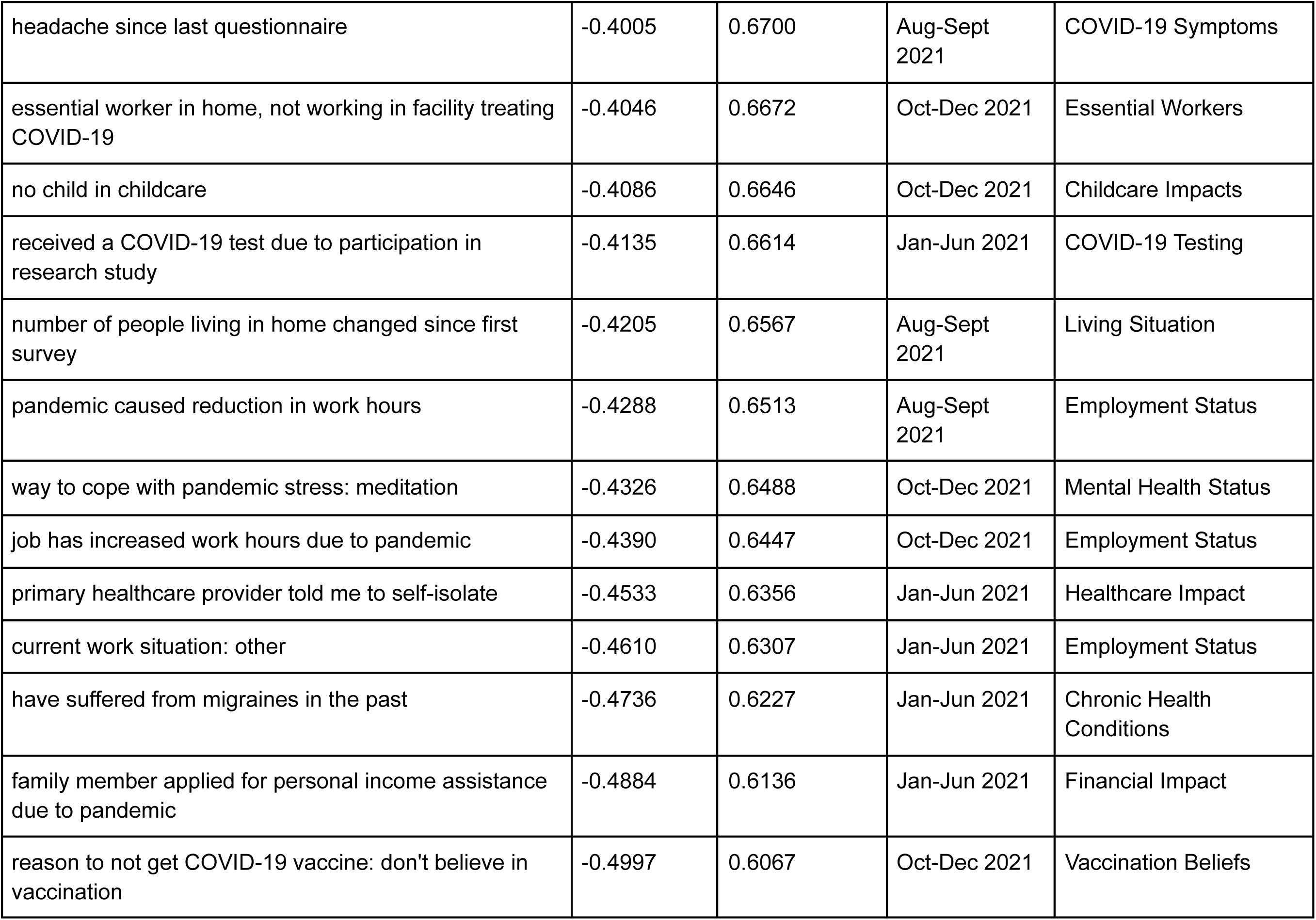
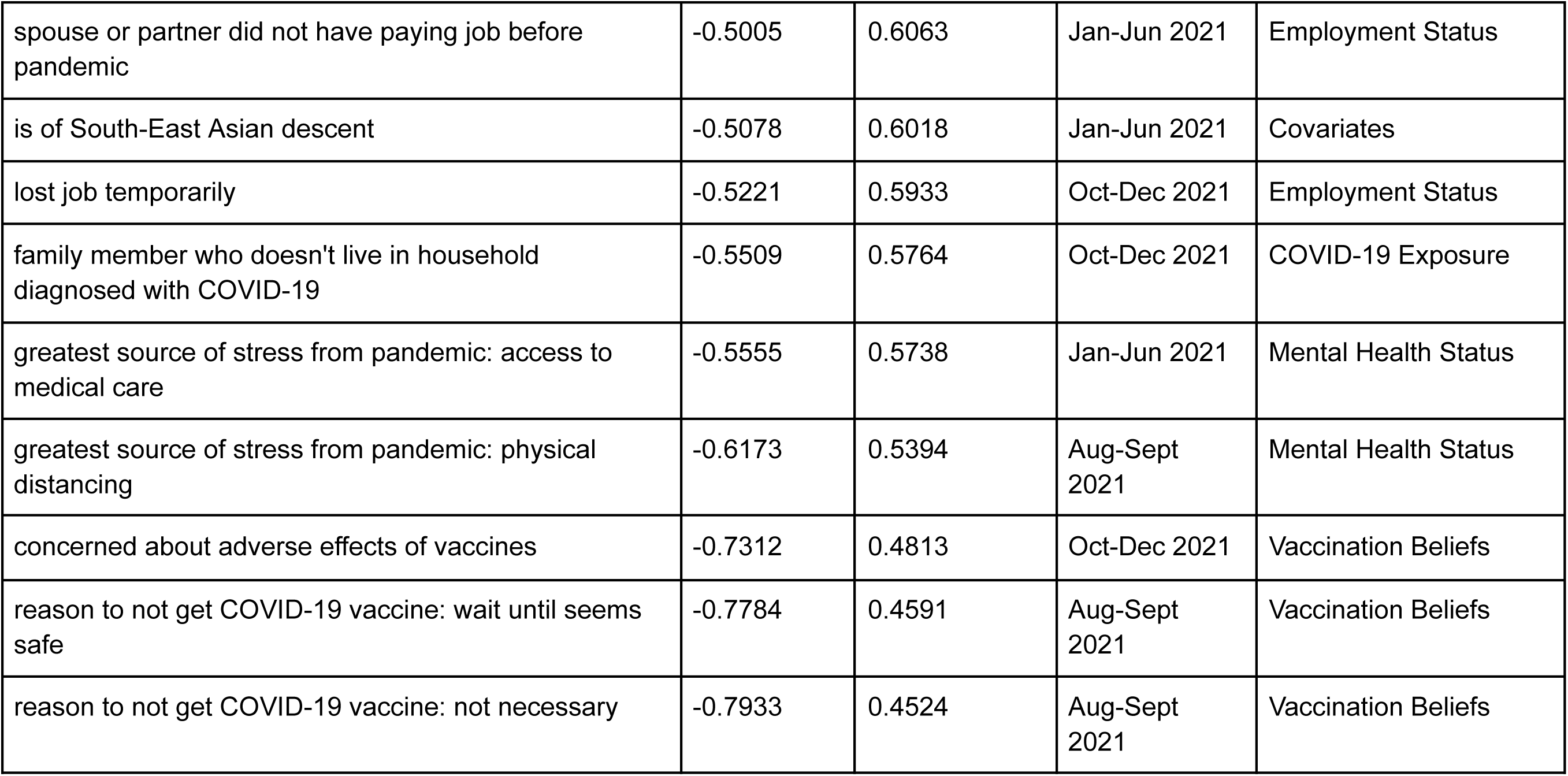
Features from modelling the response to the question “Having myself vaccinated is important for the health of others in my community” via logistic regression with scikit-learn.

**Table S4B:** Features from modelling the response to the question “Getting vaccinated is a good way to protect myself from disease’’ via logistic regression with scikit-learn.

| feature | coefficient | odds-ratio | questionnaire | domain |
| --- | --- | --- | --- | --- |
| currently staying at home / homemaker | 1.4003 | 4.0564 | Aug-Sept 2021 | Employment Status |
| source of stress from pandemic: impact on child | 1.2247 | 3.4033 | Oct-Dec 2021 | Mental Health Status |
| amount avoided crowded places | 1.1920 | 3.2937 | Jan-Jun 2021 | COVID-19 Safety Practices |
| experienced COVID-19-like symptoms, did not receive any treatment | 0.9894 | 2.6896 | Jan-Jun 2021 | COVID-19 Symptoms |
| moved to working from home | 0.9399 | 2.5597 | Jan-Jun 2021 | Employment Status |
| amount avoided crowded places/gatherings | 0.9278 | 2.5289 | Oct-Dec 2021 | COVID-19 Safety Practices |
| side effects from COVID-19 vaccine: headache | 0.9202 | 2.5098 | Aug-Sept 2021 | COVID-19 Vaccine Side Effects |
| family member applied for rent/mortgage deferral due to pandemic | 0.9129 | 2.4915 | Jan-Jun 2021 | Financial Impact |
| job has increased risk of catching COVID-19 | 0.8035 | 2.2333 | Oct-Dec 2021 | Employment Status |
| positive effects of pandemic: more exercising | 0.7962 | 2.2171 | Oct-Dec 2021 | Positive Changes |
| way to cope with stress from pandemic: drinking alcohol | 0.7790 | 2.1792 | Jan-Jun 2021 | Mental Health Status |
| experienced COVID-19-like symptoms, did not receive any treatment | 0.7167 | 2.0477 | Oct-Dec 2021 | COVID-19 Symptoms |
| had runny nose | 0.6665 | 1.9474 | Jan-Jun 2021 | COVID-19 Symptoms |
| teacher | 0.5315 | 1.7015 | Oct-Dec 2021 | Essential Workers |
| worker in a facility treating COVID-19 | 0.5309 | 1.7004 | Oct-Dec 2021 | Essential Workers |
| work in a grocery store | 0.3775 | 1.4587 | Aug-Sept 2021 | Essential Workers |
| amount avoided handshakes/hugs | 0.3638 | 1.4388 | Oct-Dec 2021 | COVID-19 Safety Practices |
| had learning disorder in the past | -0.3453 | 0.7080 | Jan-Jun 2021 | Chronic Health Conditions |
| had COVID-19-like symptoms, had contact with individual who tested positive | -0.4774 | 0.6204 | Oct-Dec 2021 | COVID-19 Exposure |
| reason to not get COVID-19 vaccine: concern about risk | -0.4880 | 0.6138 | Aug-Sept 2021 | Vaccination Beliefs |
| way to cope with pandemic stress: more reading | -0.5058 | 0.6030 | Jan-Jun 2021 | Mental Health Status |
| other adult in household is essential worker, does not come home each day: separated due to pandemic | -0.5098 | 0.6006 | Oct-Dec 2021 | Essential Workers |
| positive effects of pandemic: more exercising | -0.5674 | 0.5670 | Jan-Jun 2021 | Positive Changes |
| have had to pay more for childcare | -0.5951 | 0.5515 | Aug-Sept 2021 | Childcare Impacts |
| had irritable bowel syndrome in past | -0.5985 | 0.5497 | Jan-Jun 2021 | Chronic Health Conditions |
| amount practiced physical distancing before pandemic | -0.6202 | 0.5378 | Jan-Jun 2021 | COVID-19 Safety Practices |
| greatest source of stress from pandemic: impact on community | -0.6430 | 0.5257 | Jan-Jun 2021 | Mental Health Status |
| work in a factory | -0.6817 | 0.5058 | Oct-Dec 2021 | Essential Workers |
| work in a grocery store | -0.6896 | 0.5018 | Jan-Jun 2021 | Essential Workers |
| pandemic caused reduction in work hours | -0.8223 | 0.4394 | Aug-Sept 2021 | Employment Status |
| greatest source of stress from pandemic: impact on economy | -1.0228 | 0.3596 | Jan-Jun 2021 | Mental Health Status |
| experienced fatigue | -1.0667 | 0.3441 | Aug-Sept 2021 | COVID-19 Symptoms |
| concerned about adverse effects of vaccines | -2.5007 | 0.0820 | Oct-Dec 2021 | Vaccination Beliefs |

**Table S5A:**
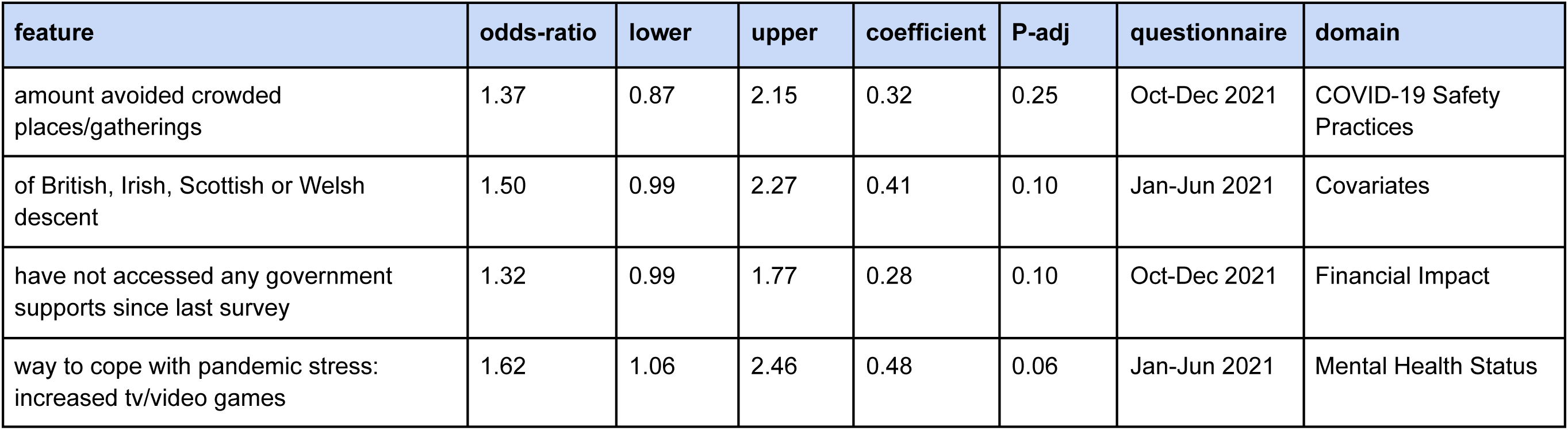
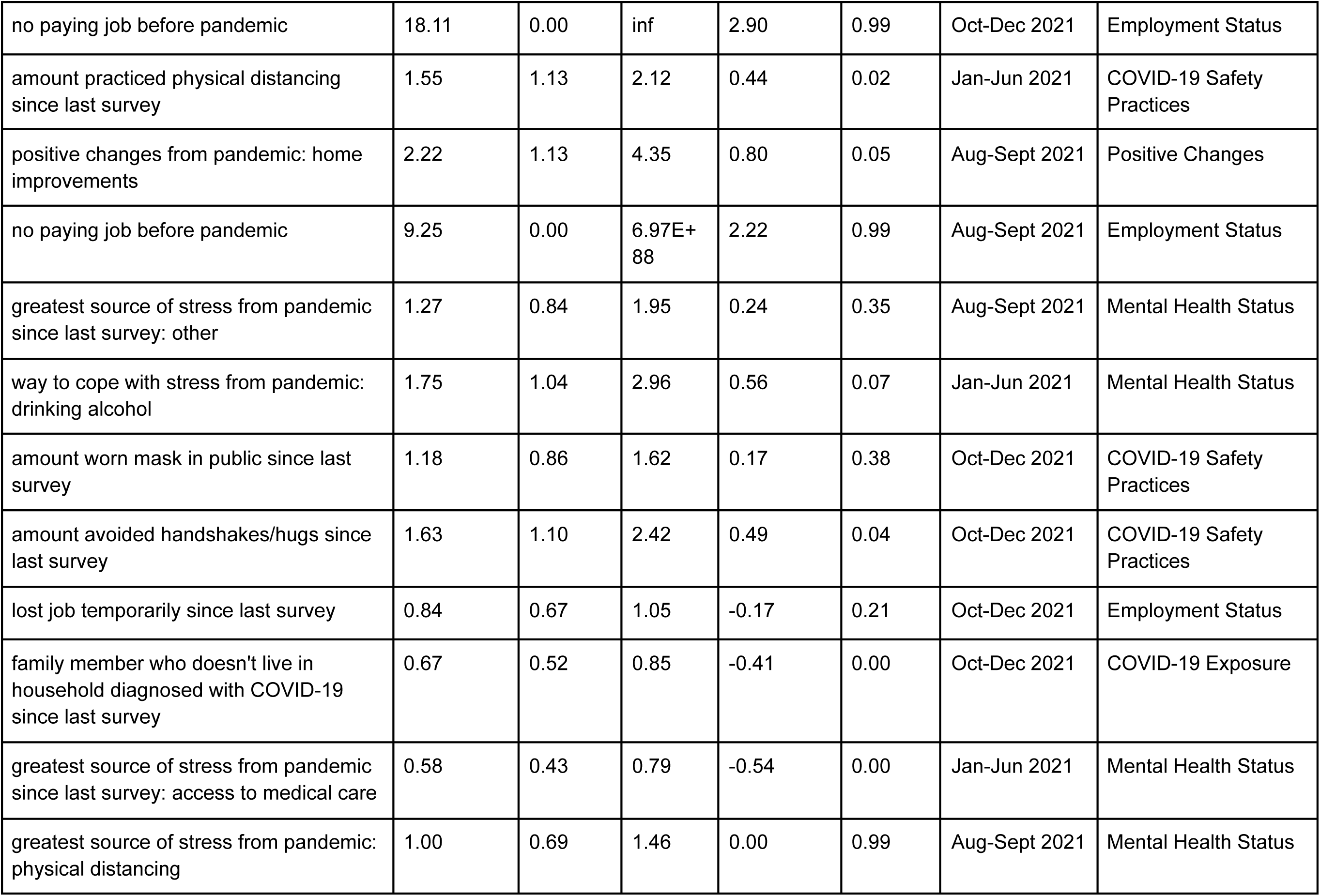
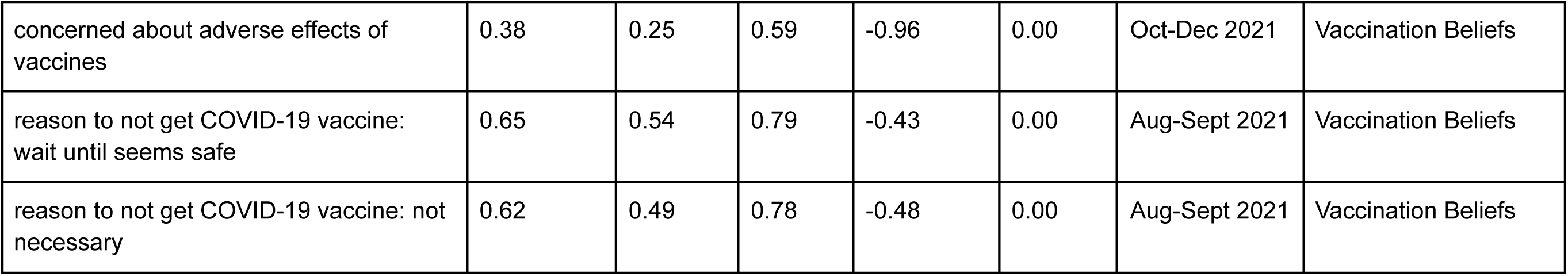
Features from modelling the response to the question “Having myself vaccinated is important for the health of others in my community” via logistic regression with statsmodels. Adjusted p-values were calculated using the Benjamini-Hochberg method.

**Table S5B:** Features from modelling the response to the question “Getting vaccinated is a good way to protect myself from disease’’ via logistic regression with statsmodels. Adjusted p-values were calculated using the Benjamini-Hochberg method.

| feature | odds-ratio | lower | upper | coefficient | P-adj | questionnaire | domain |
| --- | --- | --- | --- | --- | --- | --- | --- |
| currently staying at home / homemaker | 73117.81 | 0.00 | inf | 11.20 | 1.00 | Aug-Sept 2021 | Employment Status |
| source of stress from pandemic: impact on child | 10.83 | 1.81 | 64.88 | 2.38 | 0.04 | Oct-Dec 2021 | Mental Health Status |
| amount avoided crowded places since last survey | 8.98 | 1.85 | 43.65 | 2.19 | 0.04 | Jan-Jun 2021 | COVID-19 Safety Practices |
| experienced COVID-19-like symptoms since last survey, did not receive any treatment | 1.58 | 0.27 | 9.29 | 0.46 | 0.78 | Jan-Jun 2021 | COVID-19 Symptoms |
| moved to working from home | 18.23 | 1.45 | 229.62 | 2.90 | 0.05 | Jan-Jun 2021 | Employment Status |
| amount avoided crowded places/gatherings | 2.03 | 0.65 | 6.41 | 0.71 | 0.30 | Oct-Dec 2021 | COVID-19 Safety Practices |
| side effects from COVID-19 vaccine: headache | 6.56 | 1.18 | 36.57 | 1.88 | 0.06 | Aug-Sept 2021 | COVID-19 Vaccine Side Effects |
| family member applied for rent/mortgage deferral due to pandemic | 87287.59 | 0.00 | inf | 11.38 | 1.00 | Jan-Jun 2021 | Financial Impact |
| job has increased risk of catching COVID-19 | 8.96 | 1.53 | 52.62 | 2.19 | 0.04 | Oct-Dec 2021 | Employment Status |
| positive effects of pandemic: more exercising | 0.98 | 0.32 | 2.99 | -0.03 | 1.00 | Oct-Dec 2021 | Positive Changes |
| way to cope with stress from pandemic: drinking alcohol | 10.57 | 0.36 | 308.12 | 2.36 | 0.24 | Jan-Jun 2021 | Mental Health Status |
| experienced COVID-19-like symptoms since last survey, did not receive any treatment | 23.05 | 1.94 | 273.25 | 3.14 | 0.04 | Oct-Dec 2021 | COVID-19 Symptoms |
| had runny nose since last survey | 4.08 | 0.64 | 25.93 | 1.41 | 0.20 | Jan-Jun 2021 | COVID-19 Symptoms |
| have had to pay more for childcare since last survey | 0.50 | 0.27 | 0.92 | -0.69 | 0.05 | Aug-Sept 2021 | Childcare Impacts |
| had irritable bowel syndrome in past | 0.15 | 0.05 | 0.47 | -1.88 | 0.01 | Jan-Jun 2021 | Chronic Health Conditions |
| amount practiced physical distancing from Jan-Mar 2020 | 0.24 | 0.05 | 1.20 | -1.43 | 0.13 | Jan-Jun 2021 | COVID-19 Safety Practices |
| greatest source of stress from pandemic: impact on community | 0.22 | 0.06 | 0.84 | -1.53 | 0.05 | Jan-Jun 2021 | Mental Health Status |
| work in a factory | 0.14 | 0.00 | 5.00E+120 | -1.96 | 1.00 | Oct-Dec 2021 | Essential Workers |
| work in a grocery store | 0.69 | 0.47 | 0.99 | -0.38 | 0.08 | Jan-Jun 2021 | Essential Workers |
| pandemic caused reduction in work hours | 0.15 | 0.04 | 0.59 | -1.88 | 0.04 | Aug-Sept 2021 | Employment Status |
| greatest source of stress from pandemic: impact on economy | 0.16 | 0.04 | 0.67 | -1.82 | 0.04 | Jan-Jun 2021 | Mental Health Status |
| experienced fatigue since last survey | 0.22 | 0.07 | 0.68 | -1.53 | 0.04 | Aug-Sept 2021 | COVID-19 Symptoms |
| concerned about adverse effects of vaccines | 0.01 | 0.00 | 0.15 | -4.27 | 0.01 | Oct-Dec 2021 | Vaccination Beliefs |

